# A Comprehensive Prospective Cohort in Preventive Medicine: Protocol and Profile of the First 1,000 Participants in a Health Screening Program

**DOI:** 10.1101/2025.07.07.25330615

**Authors:** Pierre Bauvin, Alaedine Benani, Maryne Lepoittevin, Marguerite Sentilhes, Marie Bringer, Djihane Ahmed Lecheheb, Nina De Luca, Stéphane Ohayon, Claude Dalle, Xavier Tannier, Philippe Gabriel Steg, Sylvain Bodard

**Author notes:** Deceased (Claude Dalle).

## Abstract

The Zoī cohort is a prospective longitudinal cohort study, designed to advance evidence-based personalized prevention, by systematically screening for undiagnosed or asymptomatic conditions, identifying early risk markers, and predicting future disease risks. Conducted within a dedicated prevention-focused health center, data collection takes place in a standardized environment and combines over 500 self-reported items, clinical examinations, extensive biomarker profiling (196 biomarkers), and multimodal imaging (vascular, breast, abdominal, and pelvic ultrasound, full-body composition, retinal scan). For several major diseases, risk is further estimated through established clinical prediction models. Longitudinal follow-up is collected via yearly re-evaluations and through a dedicated application.

This manuscript presents the cohort design and the characteristics of the first 1,000 participants. Participants (67.5% male, mean age 51.1 years, high education levels) exhibited a high level of health awareness, lower obesity and smoking rates than the general population, yet almost half (45.6%) of those who reported no known ongoing diseases had at least one undiagnosed chronic condition (i.e., either disease or risk factor), with hypertension and hypercholesterolemia being the most frequent. Male sex and older age were significantly associated with disease unawareness (p<0.05). These findings highlight a discrepancy between self-reported and objectively measured health status, even among a well-educated and health-conscious cohort.

This deeply phenotyped, longitudinal cohort will serve as a platform that supports interdisciplinary research collaborations. It will enable development and validation of early risk stratification models, essential for predictive medicine, as well as evaluation of preventive interventions, to advance evidence-based precision prevention in public health settings.

## Why was the cohort set up?

Chronic diseases are a leading cause of morbidity and mortality in high-income countries, prompting a progressive shift from reactive to preventive healthcare models [1]. This evolution is exemplified by the Predictive, Preventive, Personalized, and Participatory (4P) medicine framework [2], which leverages advanced data analytics and individualized care strategies to anticipate health risks before the onset of disease and to support proactive health management [3,4].

In France, investment in preventive healthcare has historically been below the European average (2.8% vs. 3.2% of total health expenditure in 2020) [5,6]. This limited investment may have contributed to a sustained burden of preventable conditions such as diabetes, hypertension, obesity and lifestyle-related cancers [7]. In recent years, national health strategies have sought to reverse this trend by expanding preventive infrastructures, including digital tools and dedicated screening centers supported by both public and private initiatives, within the context of France’s universal health coverage system, which combines a single public payer with complementary private insurance schemes [8–10]. Yet, important challenges remain in embedding prevention into routine care, including inconsistent follow-up, low adherence, and variability in the quality of risk assessment and screening [11].

Several large-scale epidemiological cohorts have generated valuable insights, such as CONSTANCES in France [12]. These cohorts, while foundational, often include participants who are more educated and in better health than the general population. The Zoī cohort shares this limitation but complements existing resources by implementing a highly standardized protocol and by associating self-reported questionnaires and annual comprehensive health check-ups: all participants undergo the same assessments under controlled conditions, with minimal missing data [13,14].

Moreover, follow-up intervals are often irregular or infrequent, or based only on electronic health records, limiting the ability to detect subclinical changes or generating biased effects [15,16]. Continuous or at least yearly follow-up—ideally supported by digital tools—remains uncommon, but greatly valuable, especially when integrating multimodal data [17].

Finally, while many cohort studies have focused on primary or secondary prevention among rural or socioeconomically vulnerable populations, less attention has been paid to structured preventive health assessments among urban, health-literate individuals, especially in France [18–22]. Despite good healthcare access, these populations may still be exposed to under-recognized risk factors—such as chronic stress, air pollution, and lifestyle imbalance [23–25].

To address these gaps, the Zoī cohort was established as a prospective, open-ended study embedded in a high-throughput preventive health center in Paris. Zoī emphasizes an individualized and proactive approach. Participants undergo standardized, multimodal assessments rooted in 4P medicine—integrating clinical evaluation, imaging, advanced biomarker profiling, and continuous digital follow-up—enabling precise, actionable health insights over time.

This article describes the design, methods, and initial findings from the first 1,000 participants enrolled in the cohort. It provides insight into the population’s baseline health characteristics and highlights the potential of structured preventive assessments to detect early, undiagnosed risk conditions even in a health-aware, urban demographic.

## Who is in the cohort?

The Zoī cohort is a cohort of voluntary participants, primarily French adult residents, undergoing extensive analysis with longitudinal follow-up. Participants included in the cohort were adults aged 18 years or older, enrolled in the Zoī program either as paying customers or as beneficiaries of a company-sponsored program, since November 2023. All participants were informed prior to inclusion, with the possibility to opt out of this analysis, in accordance with national regulations. At baseline, they were invited to complete a comprehensive health and behavioral questionnaire and to attend a health screening center (HSC) in Paris, France, where they underwent a morphological, clinical, radiographic and biological assessment, performed routinely as part of the check-up. Recruitment is ongoing with no predefined end date. The Paris center is projected to have the capacity to conduct up to 2,000 check-ups per month.

A sample of the first 1,000 participants of the cohort is included in the present cross-sectional analysis. 50.4% of them were paying customers, and 49.6% are beneficiaries of a company-sponsored program. **Table 1** provides an overview of the demographic, anthropometric, and health-related characteristics of the sample. To contextualize the cohort within the broader population, comparative analyses were conducted using publicly available data from French national epidemiological cohorts [26–29], including CONSTANCES, a nation-wide health cohort [12]. We also used recent studies from the EPIC cohort for European context [30,31].

**Table 1.**
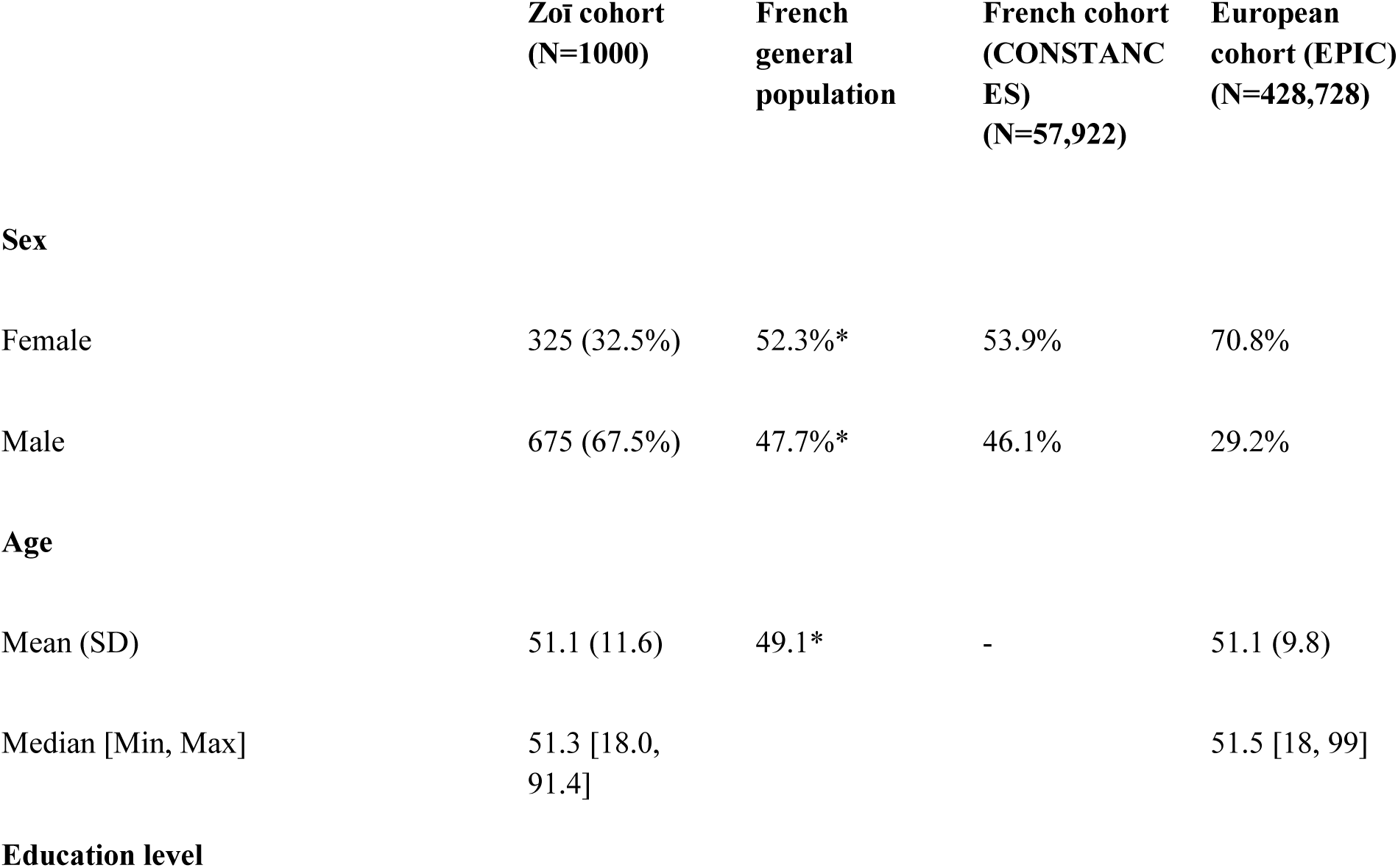
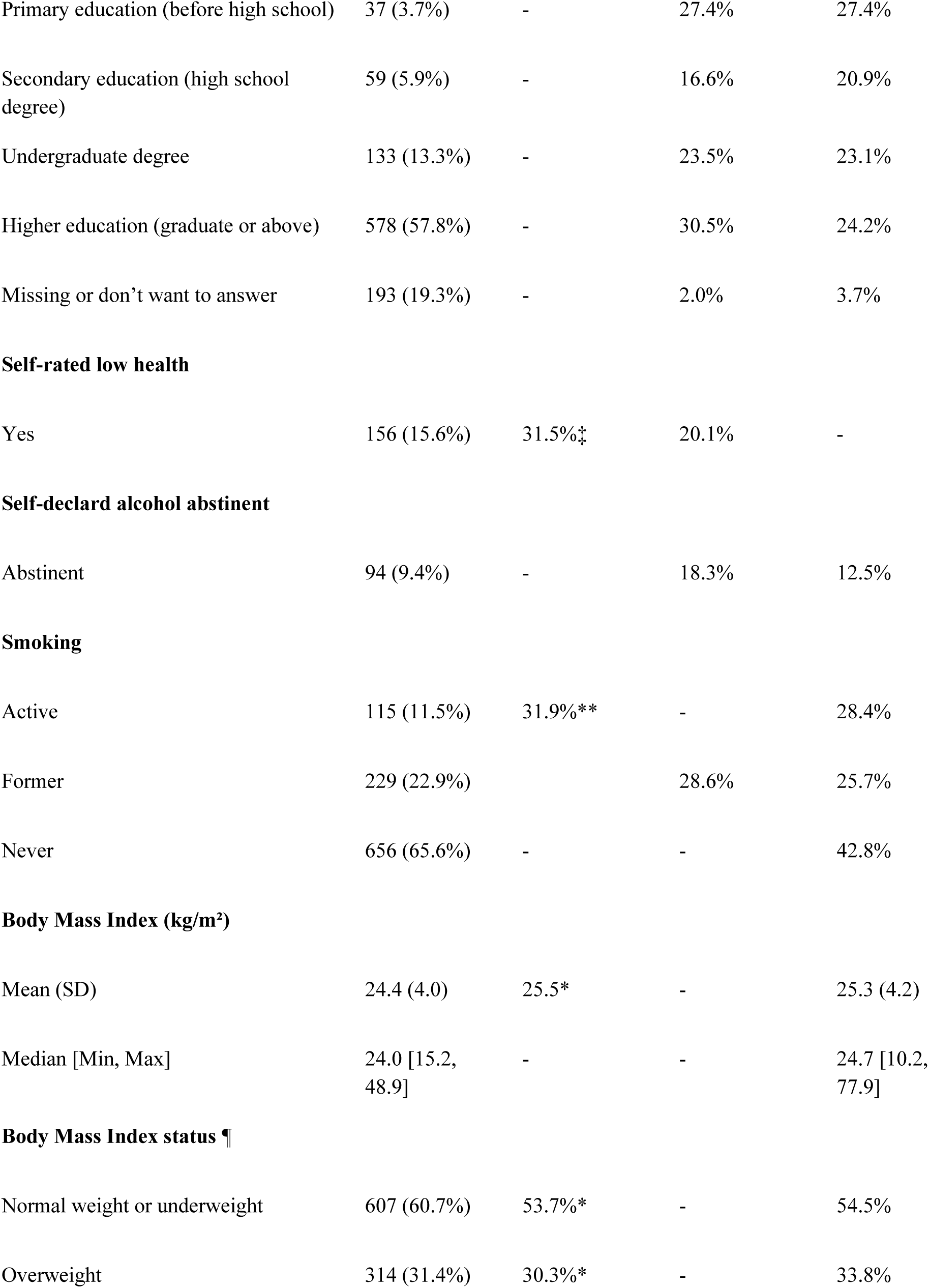
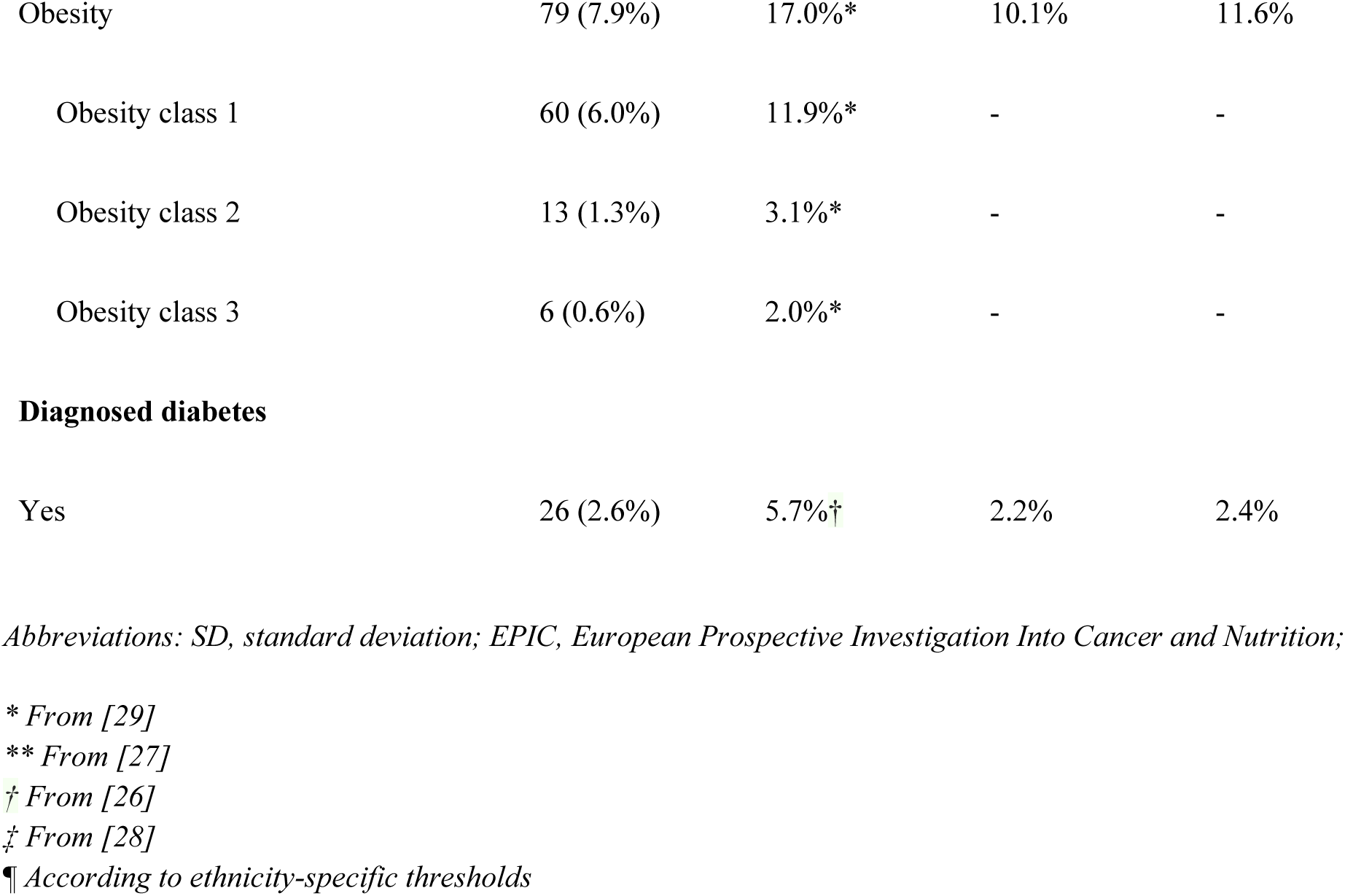
Demographic, anthropometric, and health-related characteristics of the sample with comparisons to the French general population, the CONSTANCES cohort and the European EPIC cohort.

|  | Zoī cohort<br>(N=1000) | French<br>general<br>population | French cohort<br>(CONSTANCES)<br>(N=57,922) | European<br>cohort (EPIC)<br>(N=428,728) |
| --- | --- | --- | --- | --- |
| **Sex** |  |  |  |  |
| Female | 325 (32.5%) | 52.3%* | 53.9% | 70.8% |
| Male | 675 (67.5%) | 47.7%* | 46.1% | 29.2% |
| **Age** |  |  |  |  |
| Mean (SD) | 51.1 (11.6) | 49.1* | - | 51.1 (9.8) |
| Median [Min, Max] | 51.3 [18.0,<br>91.4] |  |  | 51.5 [18, 99] |
| **Education level** |  |  |  |  |
| Primary education (before high school) | 37 (3.7%) | - | 27.4% | 27.4% |
| Secondary education (high school degree) | 59 (5.9%) | - | 16.6% | 20.9% |
| Undergraduate degree | 133 (13.3%) | - | 23.5% | 23.1% |
| Higher education (graduate or above) | 578 (57.8%) | - | 30.5% | 24.2% |
| Missing or don't want to answer | 193 (19.3%) | - | 2.0% | 3.7% |
| **Self-rated low health** |  |  |  |  |
| Yes | 156 (15.6%) | 31.5%<sup>‡</sup> | 20.1% | - |
| **Self-declared alcohol abstinent** |  |  |  |  |
| Abstinent | 94 (9.4%) | - | 18.3% | 12.5% |
| **Smoking** |  |  |  |  |
| Active | 115 (11.5%) | 31.9%** | - | 28.4% |
| Former | 229 (22.9%) |  | 28.6% | 25.7% |
| Never | 656 (65.6%) | - | - | 42.8% |
| **Body Mass Index (kg/m<sup>2</sup>)** |  |  |  |  |
| Mean (SD) | 24.4 (4.0) | 25.5* | - | 25.3 (4.2) |
| Median [Min, Max] | 24.0 [15.2, 48.9] | - | - | 24.7 [10.2, 77.9] |
| **Body Mass Index status ¶** |  |  |  |  |
| Normal weight or underweight | 607 (60.7%) | 53.7%* | - | 54.5% |
| Overweight | 314 (31.4%) | 30.3%* | - | 33.8% |
| Obesity | 79 (7.9%) | 17.0%* | 10.1% | 11.6% |
| Obesity class 1 | 60 (6.0%) | 11.9%* | - | - |
| Obesity class 2 | 13 (1.3%) | 3.1%* | - | - |
| Obesity class 3 | 6 (0.6%) | 2.0%* | - | - |
| **Diagnosed diabetes** |  |  |  |  |
| Yes | 26 (2.6%) | 5.7%† | 2.2% | 2.4% |

*Abbreviations: SD, standard deviation; EPIC, European Prospective Investigation Into Cancer and Nutrition;*
*\* From [29]*
*\*\* From [27]*
*† From [26]*
*‡ From [28]*
*¶ According to ethnicity-specific thresholds*

The cohort was predominantly male, with 325 (32.5%) women, contrasting with national data where women constitute a majority of the population (52.3% in the French general population and 53.9% in CONSTANCES). The mean age was 51.1 years (SD = 11.6, range: 18–91 years), slightly higher than that of the French general population (49.1 years) but similar to that of the EPIC cohort (51.1 years).

Regarding educational status, the cohort was highly educated, with 578 participants (57.8%) holding a graduate degree or above, compared to 30.5% in CONSTANCES. Only 3.7% reported having no education beyond primary school, substantially lower than the 27.4% observed in CONSTANCES.

Health-related factors revealed notable differences with national data. The mean BMI (Body Mass Index) was 24.4 kg/m² (SD = 4.0), slightly lower than the French general population (25.5 kg/m²). Obesity prevalence was markedly lower in the cohort (7.9%) compared to the other cohorts (French general population, 17.0%; CONSTANCES, 10.1%; EPIC, 11.6%). Diagnosed diabetes was also less frequent (2.6%) than in the general population (5.7%) but similar to the CONSTANCES and EPIC cohort (2.2% and 2.4%, respectively).

Active smoking was reported in only 11.5% of participants, considerably lower than the 31.9% prevalence reported in the French general population. Meanwhile, 22.9% were former smokers.

Self-reported health perception was relatively positive, with only 15.6% of participants rating their health as “low” or “fair,” which is notably lower than the 31.5% reported in the French general population and 20.1% in CONSTANCES. Additionally, 9.4% of participants self-declared as alcohol abstinent, which is lower than the 18.3% reported in CONSTANCES.

No differences were observed between paying customers and beneficiaries of a company-sponsored program, regarding any of the previous demographic, anthropometric, and health-related characteristics (Supplementary Table 1). Notably, no differences were observed regarding proportion of alcohol abstinent, smokers, participants with obesity or with diabetes (respectively, p=0.621, p=0.670, p=0.50 p=0.786).

## How often are participants followed up?

Following each clinical assessment, participants could receive individualized recommendations of interventions, encompassing pharmacological treatments, dietary modifications, physical activity plans, and other health-promoting behaviors, as well as therapeutic education aimed at fostering long-term self-management skills. When appropriate, referrals to medical specialists were also provided for further evaluations. Annual health reassessments are made available to participants in this cohort to track health trajectories and intervention efficacy. In between annual reassessments, participants were followed longitudinally via a dedicated smartphone application, which facilitated self-reporting at regular intervals of lifestyle factors, medication and supplement usage, and engagement in physical activities such as walking and meditation.

The application also served as a behavioral intervention tool to enhance adherence to preventive strategies. To promote sustained adherence, the application provided reminders, prompts and actionable lifestyle modifications, and employed dedicated algorithms intended to integrate the preventive measures into participants’ daily routines.

The health reassessments and follow-up data will be addressed in a subsequent study, as only the first participants have begun their follow-up visits to date.

## What is measured?

Extensive data were systematically collected from multiple sources: self-reported questionnaires, clinical examination, medical devices (electrocardiogram, spirometry), laboratory tests (urine, blood, saliva), and medical imaging, ensuring a broad assessment of health status. The protocol was designed to accommodate future scientific developments, ensuring its continued relevance and applicability.

A structured health and behavioral questionnaire was administered to all participants, encompassing sociodemographic characteristics, medical and family history, environmental exposures, dietary habits, physical activity, mental health, sleep patterns, sexual activity and other relevant domains. Standardized scales, such as the Epworth Sleepiness Scale or the STOP-BANG questionnaire, were incorporated for sleep assessment [32,33]. A detailed description of the current version of the questionnaire is provided in **Supplementary Table 2**.

During clinical examination, physical and functional parameters were recorded, including blood pressure, heart rate, heart rate variability, waist and neck circumference, and visual acuity. Systematic assessments included spirometry (Spirobank II Smart), 12-lead electrocardiography (PCA 500, QT Medical), hand grip strength testing (K-Grip, Kinvent), audiometry, and non-invasive evaluation of advanced glycation end products (AGEs) using skin autofluorescence (AGE Reader, Diagnoptics) [34].

Biological sampling included saliva, urine, and blood specimens. Saliva samples were self-collected upon awakening, 30 minutes post-awakening, and at nighttime. Urine samples included first morning void and 24-hour urine collections, while blood samples were obtained by a nurse at the HSC visit. A total of 196 biomarkers were analyzed, including standard metabolic markers and more advanced measures such as oxidative stress and neurotransmitter evaluation [35]. A full list of biomarkers is available in **Supplementary Table 3**.

Medical imaging assessments included non-invasive modalities such as vascular, breast, abdominal, and pelvic ultrasounds (ACUSON Juniper, Siemens), dual-energy X-ray absorptiometry (DEXA) for bone density and body composition (Lunar iDXA, GE Healthcare), cone beam computed tomography (CBCT) for lung, sinus, and dental assessments (NewTom 7G, NewTom), and optical coherence tomography (OCT) for retinal and optic nerve evaluation (Mirante, Nidek France) [36]. All imaging results were systematically reviewed by expert radiologists and ophthalmologists.

### Diagnoses

Diagnosed conditions encompassed both clinically diagnosed diseases and early risk markers. The latter were defined as measurable deviations from optimal health that do not meet clinical disease thresholds but may indicate an increased risk of future disease. These included: prediabetes, overweight, specific micronutrient deficiencies, fatty acid dysregulation, elevated inflammatory markers, and early sleep disturbances.

Most diagnoses were established based on clinical and biological measurements using internationally recognized criteria; e.g. diabetes and pre-diabetes were defined according to the American Diabetes Association (ADA) criteria [37], hypertension was classified based on the 2024 European Society of Cardiology (ESC) guidelines [38], hypercholesterolemia was determined using the American College of Cardiology (ACC)/American Heart Association (AHA) guidelines [35] based on cardiovascular risk profiles, obesity was derived from the recent The Lancet Diabetes & Endocrinology Commission via ethnicity-specific BMI cut-offs and excess adiposity direct measurement [40], etc. The complete list can be found in Supplementary Data. More complex diagnoses were established by MDs and extracted from medical records.

### Clinical prediction models

Several validated clinical prediction models were retrospectively applied to the cohort data, focusing on disease prevention. Selection criteria prioritized externally validated models with low risk of bias and strong applicability to the study population and measurements [41]. Risk models included cardiovascular risk assessment tools (SCORE2, SCORE2-OP, SCORE2-Diab) [42–44], and predictive models for lung (LLPv3) [45], colorectal [46], liver [47], breast cancers [48], as well as type 2 diabetes [49].

### Quality control

Standardized protocols were implemented across all clinical assessments to ensure consistency and reliability. Measurements were performed using the same equipment, with the same team of HCPs (Healthcare Professionals), under controlled environmental conditions, including lighting, sound, temperature and humidity of each room. Data collection was almost entirely automated, minimizing the risk of data entry errors and missing values. Standardized tools and classifications, including SNOMED CT, LOINC, and FHIR-like data structures, facilitated data interoperability and broader applicability of the findings [50].

For research purposes, all participant data undergo pseudonymization [51]. For free-text data, pseudonymization is performed using a machine learning approach based on a standard token classification model for general-purpose named entity recognition (NER). This model consists of a Transformer followed by multiple constrained Conditional Random Fields (CRF) classification heads [51]. To ensure confidentiality, we use a publicly available model trained on synthetic, fictitious data^1^. Identifying information is replaced with plausible alternatives. Data access is strictly restricted, and confidentiality measures adhere to the highest ethical and security standards.

The current study followed the STANDING Together consensus recommendations to tackle algorithmic bias and promote transparency, specifically the Recommendations for the Documentation of Health Datasets (**Supplementary Table 3**) [52].

## Can I get hold of the data?

The Zoī cohort is a novel resource for advancing research in predictive and preventive health, given its depth, standardization and structuration, following an open-ended design with an annual maximum capacity of tens of thousands. In alignment with the mission to democratize personalized medicine, the Zoī cohort welcomes scientific collaborations with research projects that align with the commitment to understand, develop and implement preventive and predictive healthcare strategies. Access to the Zoī cohort can only be provided at no cost to investigators, ensuring that financial barriers do not hinder scientific progress.

Hence, collaboration with Zoī is guided by strict adherence to legal and ethical standards, prioritizing data confidentiality and security, with rigorously validated tools that preclude the possibility of individual re-identification [53]. Proposals for research projects involving the Zoī cohort are to be reviewed by the Zoī Scientific and Medical Committee that evaluates missions based on scientific merit, and an independent Ethical Committee for ethical considerations.

## What has it found? Insights from the first 1,000 participants

### Biological measurements

Key biological parameters were assessed in the cohort, providing insights into renal function, lipid metabolism, and metabolic health.

As examples, the LDL cholesterol, the Glomerular Filtration Rate (GFR), uric acid levels, and apolipoprotein B are displayed in Figure 1.

**Fig. 1:**
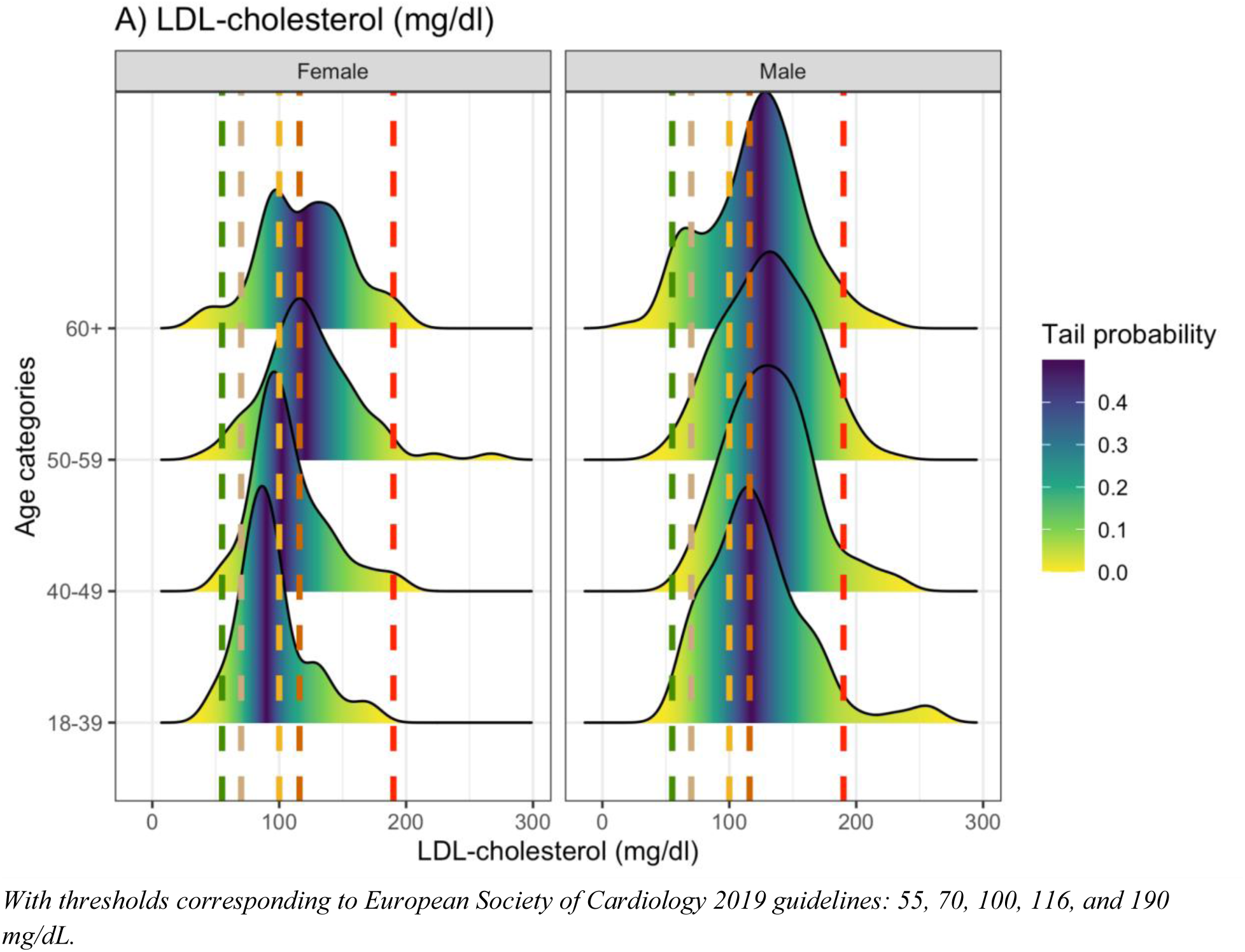
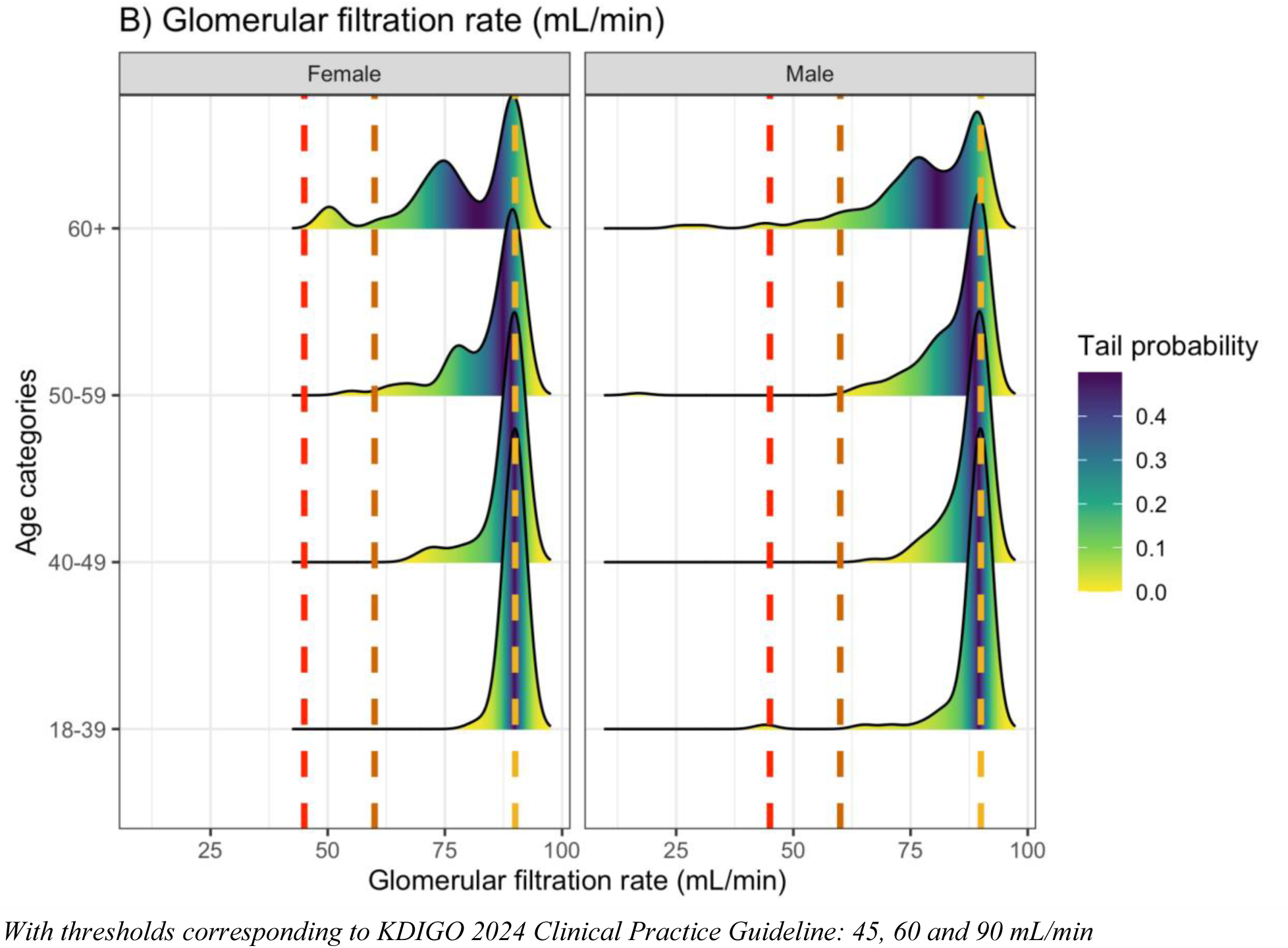
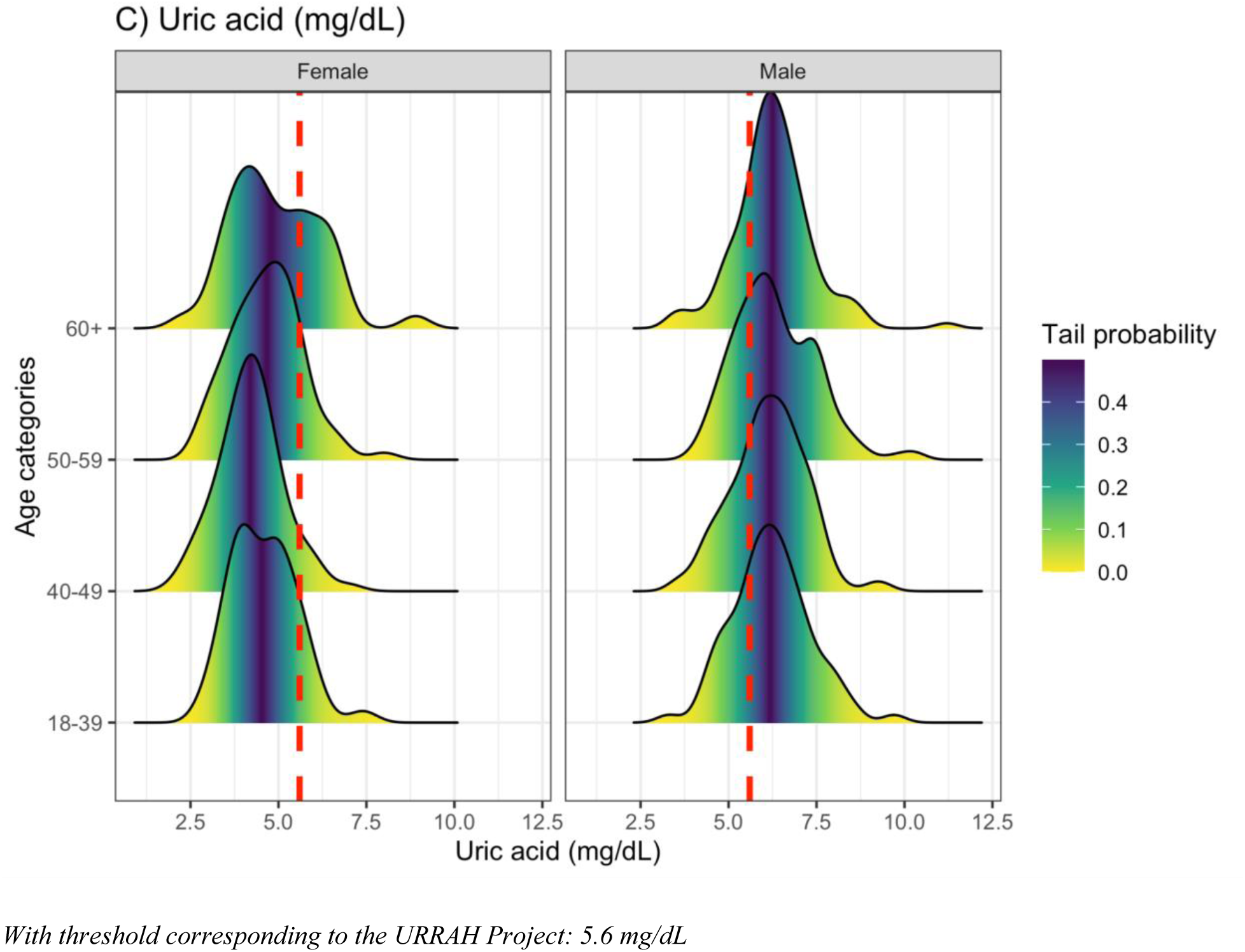
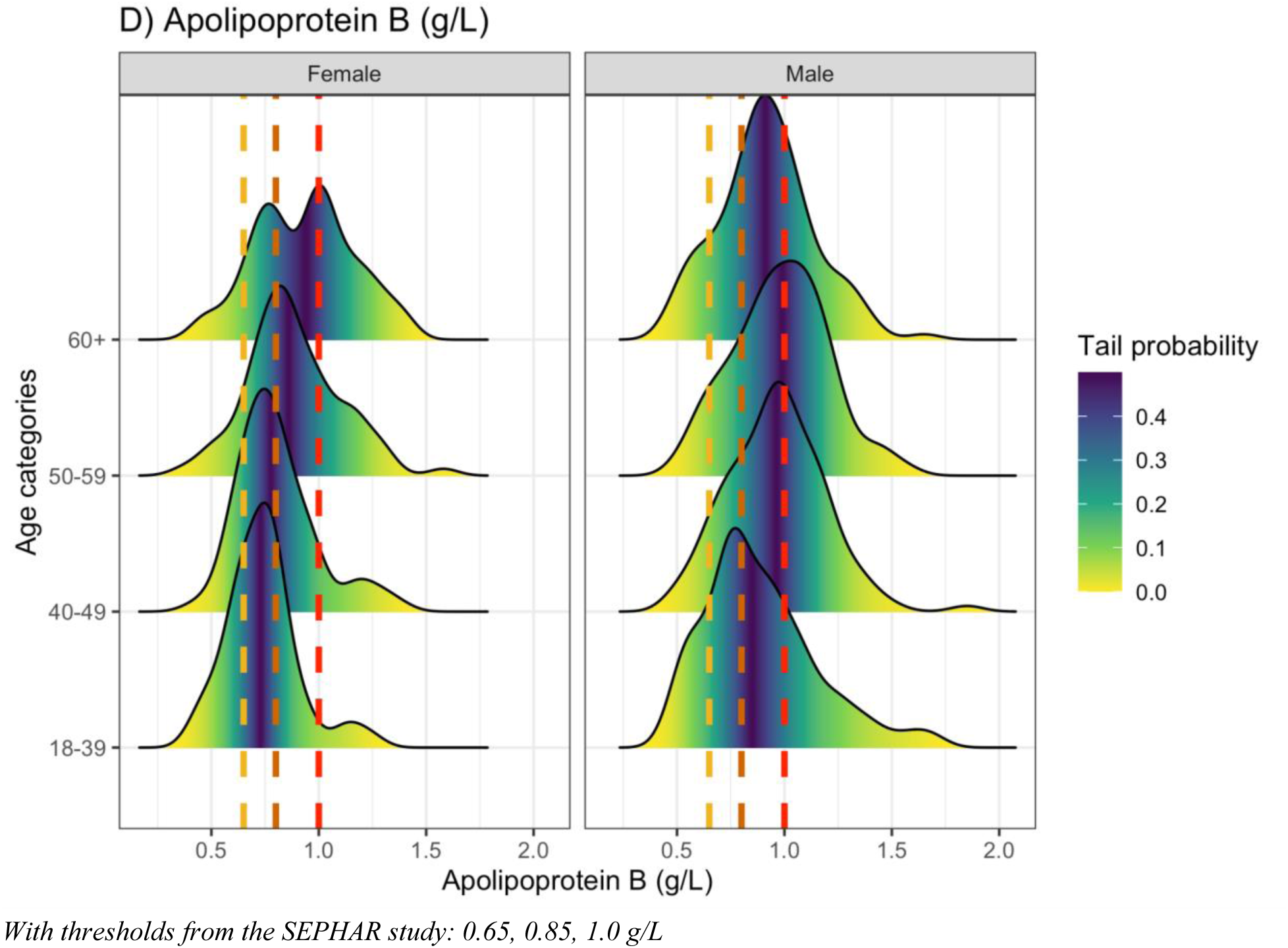
Distribution of some specific health parameters measured in the sample, by age and sex: (A) LDL-cholesterol, (B) glomerular filtration rate, (C) uric acid, (D) Apolipoprotein B.

The mean LDL-cholesterol was 122.3 (±37) mg/dL, with a majority of participants, especially males, exceeding the European Society of Cardiology 2019 thresholds: 98.5% are above 55 mg/dL, 93.3% above 70 mg/dL and 68.8% are above 100 mg/dL (**Fig. 1A**) [54].

The mean GFR was 84.8 ± 9 mL/min, with 44.8% of participants below 90 mL/min, 2.2% below 60 mL/min and 0.7% below 45 mL/min, the KDIGO 2024 guidelines thresholds [55]. Age-stratified distributions indicated a progressive decline with age (**Fig. 1B**).

Uric acid levels showed a mean of 5.7 ± 1 mg/dL, with a substantial proportion (52.4%) of participants exceeding the URRAH Project threshold of 5.6 mg/dL, particularly in males and older age groups (**Fig. 1C**) [56].

Apolipoprotein B (ApoB) had a mean concentration of 0.9 ± 0.2 g/L, with distributions indicating a shift towards higher values in men compared to women. Reference thresholds of 0.65, 0.85, and 1.0 g/L from the SEPHAR study were used for comparison (**Fig. 1D**) [57].

Additional measurements of interest, associated with cardiovascular and metabolic health, include Apolipoprotein A1 (mean 1.5 ± 0.2 g/L), fasting blood sugar (mean 87.5 ± 14.7 mg/dL), HbA1c (mean 5.3 ± 0.5 %), HOMA index (mean 1.2 ± 0.9), lipoprotein A (mean 25.3 ± 23.2, median 13.0 mg/dL). The complete list of all biomarkers is available in **Supplementary Table 2**.

### Prevalence of conditions and comparison with self-reported health status

Almost all participants (n=905, 90.5%) were found to have at least one disease or early risk marker during the comprehensive health check-up, known beforehand or not: 56.9% with at least one disease, and 88.2% with at least one early risk marker. The most common conditions were two early risk markers: fatty acid dysregulation (69.8%) and iodine deficiency (61.6%). For comparison, hypertension was found in 22.6% of the participants, known beforehand or not.

Participants were asked if they had a known ongoing diagnosis of some chronic conditions: diabetes, obesity, hypertension, hypercholesterolemia, osteoporosis, renal deficiency, hyperthyroidism, sleep apnea, or MASLD (metabolic dysfunction-associated steatotic liver disease). Among participants who self-reported having no known such condition (n = 539), only 54.4% were confirmed as having none of these diagnoses upon clinical evaluation. The remaining 45.6% were found to have at least one previously undiagnosed chronic condition: 164 (30.4%) had one condition, 70 (13.0%) had two conditions, 11 (2.0%) had three conditions, and 1 (0.2%) had four conditions (**Table 2B**).

**Table 2.** Prevalence of chronic conditions in the cohort, (A) presence or absence of diagnoses among members specifically declaring the absence of said chronic condition, (B) number of chronic conditions measured among members self-reporting as healthy.

**A) Presence or absence of diagnoses among members declaring the absence of said chronic condition**
| **Chronic condition declared not diagnosed by participants\*** | **Confirmed absent** | **Identified during check-up** |
| --- | --- | --- |
| Hypercholesterolemia (from ACC/AHA guidelines based on cardiovascular risk profiles) | 220 (48.6%) | 233 (51.4%) |
| Hypertension | 491 (78.7%) | 133 (21.3%) |
| Prediabetes | 487 (86.4%) | 78 (13.6%) |
| MASLD | 707 (94.0%) | 45 (6.0%) |
| Sleep apnea syndrome | 306 (95.0%) | 16 (5.0%) |
| Osteoporosis | 495 (98.0%) | 10 (2.0%) |
| Obesity | 681 (98.1%) | 13 (1.9%) |
| Diabetes | 565 (98.4%) | 9 (1.6%) |
| Renal deficiency | 505 (98.6%) | 7 (1.4%) |
| Biological hyperthyroidism | 539 (99.6%) | 2 (0.4%) |
| Declaring none of the above | 293 (54.4%) | 246 (45.6%) |

*Abbreviation: MASLD, Metabolic dysfunction-Associated Steatotic Liver Disease*
*\* Patients not responding to a question about the previous diagnosis of a disease were excluded from the corresponding analysis, to ensure that we only consider patients actively declaring no disease.*

**B) Number of chronic conditions identified among participants self reporting as healthy**
| Number of actual conditions | Count |
| --- | --- |
| 0 | 293 (54.4%) |
| 1 | 164 (30.4%) |
| 2 | 70 (13.0%) |
| 3 | 11 (2.0%) |
| 4 | 1 (0.2%) |

Hypercholesterolemia (defined from ACC/AHA guidelines based on cardiovascular risk profiles) was frequently underdiagnosed, with 51.4% of participants who reported no prior diagnosis of hypercholesterolemia being newly identified with the condition (**Table 2B**). Hypertension was newly diagnosed in 21.3% of participants who had previously reported no history of the condition, distributed as follows: 17.9% in stage 1, 2.9% in stage 2, and 0.5% in stage 3 (**Fig. 2**).

**Fig. 2:**
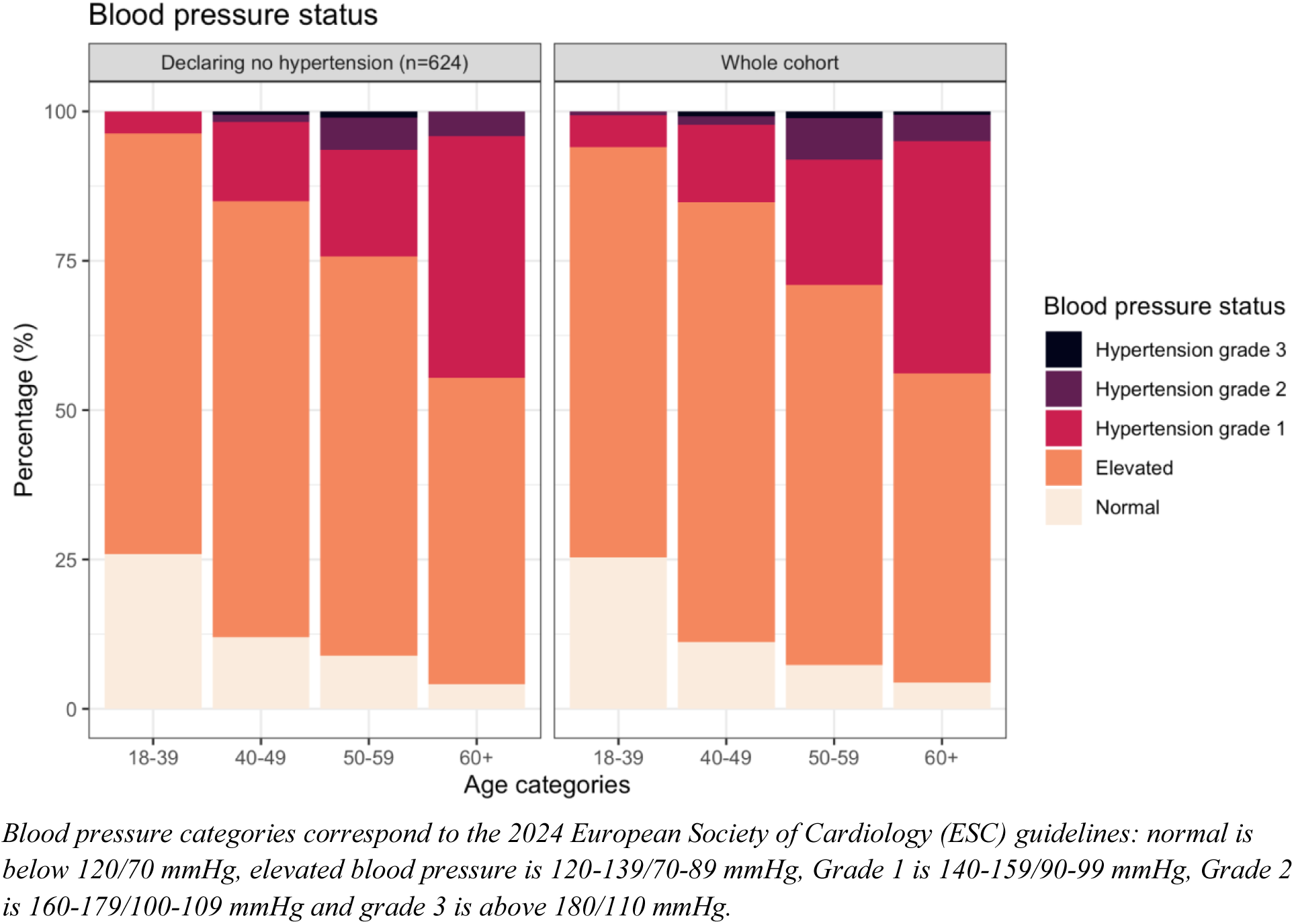
Distribution of blood pressure status in the total population, and in the subgroup self-declaring no hypertension

In a multivariable regression analysis assessing factors associated with unawareness of a current condition, while adjusting for potential confounders (age, sex, income level, and self-perceived health status), higher education levels were not significantly associated with unawareness of a current diagnosis (p = 0.605). However, male sex (p < 1e-5) and older age (1 year increment, p < 1e-5) were strongly associated with unawareness of condition.

### Identification of suspicious lesions

A total of 136 patients (13.6%) were prescribed additional investigations to further characterize or monitor a potentially tumoral lesion. These complementary explorations were recommended based on initial imaging or biology findings or clinical suspicion. Follow-up data and final diagnostic outcomes will be the subject of a subsequent study.

## What are the main strengths and weaknesses?

The Zoī cohort study presents several key strengths. Its comprehensive and standardized health assessment protocol enables detailed characterization of participants’ health status. This includes an extensive array of data sources— self-reported questionnaires, clinical examinations, medical devices, laboratory analyses, imaging techniques, and follow-up measures—collected under controlled conditions. The analysis of a first sample has revealed a substantial mismatch between perceived and actual health status: among participants reporting no prior diagnoses, 45.6% were found to have at least one common condition during the check-up. This underscores the potential of structured assessments to uncover undiagnosed disease, even in populations with high access to healthcare.

Moreover, it offers valuable opportunities for machine learning applications in the health domain. While datasets derived directly from routine care are often larger and may better represent the general population, they are typically collected for clinical purposes. This difference in intent introduces distinct types of bias, such as selection bias due to care pathways or missing data patterns influenced by clinical practices. In contrast, the dataset described here is comprehensive in both its collection and processing, ensuring higher data quality. This completeness significantly enhances its utility for machine learning tasks, as models can be trained without requiring complex imputation strategies or introducing additional noise. Furthermore, the dataset’s well-defined structure and consistent annotations facilitate the development of robust algorithms, making it particularly suited for benchmarking and model evaluation, beyond the simple bigger-is-better paradigm [58].

Another strength lies in the early detection of subclinical risk states. The assessment of early risk markers has proven to be a key component in identifying individuals at heightened risk for future disease onset [59], suggesting that even in the absence of overt disease, early deviations from optimal health may be detectable. Additionally, integrating a wide range of validated predictive scores could further refine risk stratification and inform predictive medicine strategies [60,61].

Additional strengths of the study include: dataset’s completeness and quality control, longitudinal data collection, enabling the evaluation of predictive tools or preventive strategies, and adaptability of the protocol to incorporate future scientific advances.

However, the cohort has several limitations. The study population is predominantly middle-aged and highly educated, with lower smoking and obesity prevalence and better self-rated health than the general French population. While this may limit generalizability, it provides a valuable opportunity to examine health trajectories in a health-conscious subpopulation presumed to be well-informed and with high access to healthcare services.

Another limitation concerns the current state of the protocol regarding diseases and early risk markers assessment. Most diagnoses were derived from clinical and biological measurements using internationally recognized criteria. While this approach enables standardisation and ensures up-to-date diagnostic classification, it isn’t suitable for more complex diseases that require access to detailed medical records. As a result, some diagnoses may be missed, or may necessitate manual, retrospective assessments by trained clinicians. Efforts are currently underway to improve the protocol by incorporating systematic disease reporting during check-ups by the team of HCPs, using the ICD-10 classification. This enhancement aims to reduce the number of missed diagnoses and lessen the reliance on retrospective evaluations.

As a longitudinal, deeply phenotyped cohort with ongoing data collection, the Zoī cohort is well-positioned to contribute to the evolving landscape of predictive medicine. Its design supports future collaborations, risk model development and evaluation, and policy-relevant insights into early intervention effectiveness.

## Data Availability

All data produced in the present study are available upon reasonable request to the authors

## Acknowledgments

The Zoī Preventive Medicine, Data Science and AI Lab is funded by Zoī SAS, Paris, France. The authors also express their thanks to Kévin David-Girard, Marion De Meslon, Damien Grosgeorge, Nassim Hmidou, Alexandre Hollard, Paul Roujansky, Léon-Marie de Souza, and Élodie Vorkaufer.

We pay tribute to Dr. Claude Dalle, whose vision and commitment were foundational to this protocol. A pioneer in the field of preventive medicine, he profoundly shaped the scientific thinking that led to this work.

## Statements & Declarations

### Author Contributions

Pierre Bauvin, Alaedine Benani, Marguerite Sentilhes, Xavier Tannier, Claude Dalle, Eric Vibert, Philippe Gabriel Steg, Sylvain Bodard contributed to the study conception and design. Material preparation, data collection, and analysis were performed by Pierre Bauvin, Alaedine Benani, Marie Bringer, Nina De Luca, Stéphane Ohayon. The first draft of the manuscript was written by Pierre Bauvin, Djihane Ahmed Lecheheb and Sylvain Bodard, and all authors reviewed and approved the final manuscript, except Claude Dalle who passed away before the completion of the final draft.

### Ethics approval

The study was approved by the Ethics Committee of the Adène Research Foundation (IRB accreditation no.0990-0279)

### Consent to participate

The study involved only retrospective, pseudonymized data. All participants were informed generally and individually prior to inclusion, with the possibility to object, in accordance with national regulations.

### Consent to publish

No individual-identifiable data or images are included in this manuscript.

## Footnotes

1 https://github.com/aphp/eds-pseudo

