## Supplemental File for "A Comprehensive Prospective Cohort in Preventive Medicine: Protocol and Profile of the First 1,000 Participants in a Health Screening Program"

### Supplementary Information - Statistical Analyses and Health Risk Stratification in the First 1,000 Participants

#### Statistical analysis

Descriptive statistics were used to summarize the demographic and health-related characteristics of the cohort. Continuous variables were expressed as means with standard deviations (SD), while categorical variables were presented as proportions.

Multivariable logistic regression models were employed to assess associations between demographic characteristics, health behaviors, and clinical outcomes, adjusting for potential confounders. Results were expressed as odds ratios (OR) with 95% confidence intervals (CI). A two-sided significance threshold of p < 0.05 was applied. Missing data were handled using pairwise deletion to ensure minimal impact on the overall analysis. All statistical analyses were conducted using R software version 4.4.0 .

#### Health risk scores

Several validated risk scores were calculated at the time of the health check-up to estimate the likelihood of future events. **Suppl. Fig 1** displays three scores concerning cardiovascular events, liver cancer, and lung cancer as examples.

The SCORE2 algorithm was used to assess the 10 years risk of fatal and non-fatal major cardiovascular events (stroke and myocardial infarction), using SCORE2-OP or SCORE2-Diab when relevant. A significant sex-based difference was observed, with the majority of women classified as low risk, while most men fell into the medium-risk category. A non-negligible proportion (26.5%) of men were classified as high risk, highlighting a greater cardiovascular burden in this group (**Suppl** **Fig. 1A**).

The aMAP score**,** which evaluates the risk of liver cancer, indicated that the vast majority of women had a low-risk profile. In contrast, men exhibited a more even distribution between low and medium risk, with a small subset classified as high risk (6.9%) (**Suppl Fig. 1B**).

The LLPv3 score, used for lung cancer risk assessment, showed that most participants fell into the low-risk category, with very few individuals classified as high risk (0.9%) (**Suppl Fig. 1C**).

#### Diseases

More than half of the participants were diagnosed with a disease (56.9%). For example, 22.6% of participants were diagnosed with hypertension, 46.3% of participants were diagnosed with hypercholesterolemia (from ACC/AHA guidelines based on cardiovascular risk profiles), and 2.6% of participants were diagnosed with diabetes. 7.9% of participants were diagnosed with clinical obesity, according to BMI category and excess adiposity.

Overall, 431 members were not diagnosed with a disease, whereas 323 (32.3%) were diagnosed with 1 disease, 171 (17.1%) with 2 diseases, 59 (5.9%) with 3 diseases, and 16 (1.6%) with 4 or more diseases simultaneously.

#### Early risk markers

Early risk markers were also identified, with 88.2% of participants exhibiting at least one such marker and 31.8% presenting with three or more. The most common early risk markers were fatty acid dysregulation (69.8%) and iodine deficiency (61.6%).

Regarding cardiovascular and metabolic health, a total of 28.7% of participants were identified with hyperhomocysteinemia, 39.3% with overweight, and 10.4% with prediabetes.

### The main factor associated with the presence of early risk markers, independently of the presence of diagnosed diseases, was younger age (p < 1e-5). This association was identified through a multivariable regression analysis restricted to participants without any diagnosed disease (n = 384) and adjusted for potential confounders. However, longitudinal follow-up is not yet available for this cohort, and will be part of future investigation to assess the long-term implications of these findings. Definition of diseases and risk markers referenced in the manuscript

Body mass index (BMI) status was determined according to the World Health Organization (WHO) definitions, with modifications applied for individuals of Asian and Pacific ethnicities, in accordance with established guidelines (1). For participants of non-Asian ethnicity, BMI values <25.0 kg/m² were classified as normal weight or underweight, values between 25.0 and 29.9 kg/m² as overweight, values between 30.0 and 34.9 kg/m² as obesity class I, values between 35 and 39.9 as obesity class II, and values ≥40.0 kg/m² as obesity class III. For participants identified as being of Asian or Pacific ethnicity, lower thresholds were applied to account for differential health risk profiles. In this group, BMI values <23.0 kg/m² were classified as normal weight, values from 23.0 to 25.0 kg/m² as overweight, and values ≥25.0 kg/m² as obesity. Ethnicity was determined based on self-reported ancestry.

The diagnosis of clinical obesity was defined following a recent review as a composite criteria of BMI category (“obesity” categories) and a measure of excess adiposity by direct measurement of body fat via DEXA: a percentage body fat above 25% for males, and above 30% for females (2).

Diabetes was defined following the ADA criteria (3) by presence of antidiabetic treatment, or HbA1c above 6.5%, or fasting plasma glucose above 126 mg/dL. Prediabetes was similarly defined by HbA1c between 5.7% and 6.5%, or fasting plasma glucose between 100 mg/dL and 126 mg/dL.

Hypertension was defined following the ESC 2024 guidelines (4) by presence of hypertensive treatments, or systolic blood pressure above 140 mmHg, or diastolic blood pressure above 90 mmHg. The different hypertension categories are defined similarly: systolic blood pressure above 180 mmHg, or diastolic blood pressure above 110 mmHg were classified as hypertension grade 3, systolic blood pressure between 160 and 179 mmHg or diastolic blood pressure between 100 and 109 mmHg was classified as grade 2 hypertension; and systolic blood pressure between 140 and 159 mmHg or diastolic blood pressure between 90 and 99 mmHg was classified as grade 1 hypertension. Moreover, elevated blood pressure was defined as systolic blood pressure between 120 and 139 mmHg and/or diastolic blood pressure between 70 and 89 mmHg, whereas normal blood pressure corresponds to systolic blood pressure below 120 or diastolic blood pressure below 70.

Hypercholesterolemia was defined following the ESC/EAS guidelines (5) by presence of treatment of hypercholesterolaemia, or low-density lipoprotein-cholesterol (LDL-C) levels depending on the risk profile: LDL-C ≥190 mg/dL, or LDL-C ≥155 mg/dL and a cardiovascular risk value ≥1%, or LDL-C ≥70 mg/dL and a cardiovascular risk value ≥5%.

Osteoporosis was defined from bone densitometry measured via DEXA, following the WHO criteria (6) as a bone mineral density (BMD) that lies 2.5 standard deviation (SD) or more below the average value (T-score < − 2.5 SD). Osteopenia was similarly defined by T-score < − 1.0 SD.

Renal deficiency was defined following the KDIGO 2024 Guidelines (7) by eGFR (glomerular filtration rate by creatinine-based formula) below 60 mL/min.

Metabolic dysfunction-associated steatotic liver disease (MASLD) was suspected by either a high FIB4 score (9), a high NAFLD fibrosis score (9) or a high Fatty Liver Index (FLI) score (8). T(10)e scoring systems incorporate two distinct thresholds: an upper threshold, above which the presence of disease (liver fibrosis or steatosis) is considered likely, and a lower threshold, below which the disease can be reasonably ruled out. In the present study, the upper thresholds were applied to identify individuals with a suspected diagnosis of metabolic dysfunction-associated steatotic liver disease (MASLD). Following the original publications, we used the following high thresholds: a FIB4 value above 2.67; a NAFLD Fibrosis score above 0.676; a FLI above 60.

Sleep apnea syndrome was suspected following the STOP-BANG score for obstructive sleep apnea (11). We used a threshold corresponding to a Canadian external validation study of the score: ≥5 (12).

Fatty acid dysregulation is defined as the presence of out of range values (using the lab standard ranges) from a set of multiple biomarkers related to fatty acid regulation : omega 3 index, ratio of omega 6 (w6) on omega 3 (w3) fatty acids, ratio of omega 3 on total fatty acids, ratio of omega 6 on total fatty acids, ratio of stearic acid on total fatty acids, ratio of elaidic acid on total fatty acids, ratio of eicosatrienoic acid on total fatty acids, ratio of alpha-linolenate on total fatty acids, ratio of arachidonate on total fatty acids, ratio of trans acid CLA2 C18:2(10t,12c) on total fatty acids, ratio of trans acid CLA3 C18:2(9c,12t) on total fatty acids, ratio of eicosapentaenoate on total fatty acids, ratio of linoleate on total fatty acids, ratio of oleate on total fatty acids, ratio of palmitate on total fatty acids, ratio of palmitoleate on total fatty acids, ratio of rumenate on total fatty acids, ratio of trans-palmitoleate on total fatty acids, ratio of trans-vaccenate on total fatty acids, ratio of vaccenate on total fatty acids, ratio of arachidonate on eicosapentaenoate acid, ratio of linoleate on dihomo-gamma-linoleneate.

Iodine deficiency was defined following the WHO criteria for iodine deficiency as iodine ≤100 µg/L (13).

Vitamin D deficiency was defined as 25-hydroxyvitamin D3+D2 ≤30 µg/L following (14).

Hyperhomocysteinemia was defined as homocysteine ≥15 µmol/L following (15).

Hyperthyroidism was defined as thyroid-stimulating hormone (TSH) too low, and free thyroxine (T4) too high or free triiodothyronine (T3) too high, using standard laboratory ranges, following (16).

### Supplementary Table 1 - Demographic, anthropometric, and health-related characteristics of the sample with comparisons between paying customers and beneficiaries of a company-sponsored program

|  | **Zoī cohort (N=1000)** | **Beneficiaries of a company-sponsored program (N=496)** | **Paying customers (N=504)** | **P-value*** |
| --- | --- | --- | --- | --- |
| **Sex** |  |  |  |  |
| Female | 325 (32.5%) | 143 (28.8%) | 182 (36.1%) | 0.167 |
| Male | 675 (67.5%) | 353 (71.2%) | 322 (63.9%) |  |
| **Age** |  |  |  |  |
| Mean (SD) | 51.1 (11.6) | 51.4 (10.7) | *50.8 (12.4)* | 0.650 |
| Median [Min, Max] | 51.3 [18.0, 91.4] | 51.4 [18.0, 91.4] | 50.4 [21.0, 87.2] |  |
| **Education level** |  |  |  |  |
| Primary education (before high school) | 37 (3.7%) | 21 (4.2%) | 16 (3.2%) | 0.758 |
| Secondary education (high school degree) | 59 (5.9%) | 28 (5.6%) | 31 (6.2%) |  |
| Undergraduate degree | 133 (13.3%) | 56 (11.3%) | 77 (15.3%) |  |
| Higher education (graduate or above) | 578 (57.8%) | 285 (57.5%) | 293 (58.1%) |  |
| Missing or don’t want to answer | 193 (19.3%) | 106 (21.4%) | 87 (17.3%) |  |
| **Self-rated low health** |  |  |  |  |
| Yes | 156 (15.6%) | 61 (12.3%) | 95 (18.8%) | 0.103 |
| **Self-declard alcohol abstinent** |  |  |  |  |
| Abstinent | 94 (9.4%) | 42 (8.5%) | 52 (10.3%) | 0.621 |
| **Smoking** |  |  |  |  |
| Active | 115 (11.5%) | 61 (12.3%) | 54 (10.7%) | 0.670 |
| Former | 229 (22.9%) | 104 (21.0%) | 125 (24.8%) |  |
| Never | 656 (65.6%) | 331 (66.7%) | 325 (64.5%) |  |
| **Body Mass Index (kg/m²)** |  |  |  |  |
| Mean (SD) | 24.4 (4.0) | 24.6 (3.84) | 24.2 (4.19) | 0.456 |
| Median [Min, Max] | 24.0 [15.2, 48.9] | 24.1 [15.9, 45.4] | 23.9 [15.2, 48.9] |  |
| **Weight status** |  |  |  |  |
| Normal weight or underweight | 607 (60.7%) | 288 (58.1%) | 319 (63.3%) | 0.648 |
| Overweight | 314 (31.4%) | 163 (32.9%) | 151 (30.0%) |  |
| Obesity | 79 (7.9%) | 45 (9.1%) | 34 (6.8%) |  |
| Obesity class 1 | 60 (6.0%) | 38 (7.7%) | 22 (4.4%) |  |
| Obesity class 2 | 13 (1.3%) | 5 (1.0%) | 8 (1.6%) |  |
| Obesity class 3 | 6 (0.6%) | 2 (0.4%) | 4 (0.8%) |  |
| **Diagnosed diabetes** |  |  |  |  |
| Yes | 26 (2.6%) | 11 (2.2%) | 15 (3.0%) | 0.786 |

* The p-value calculations are based on one-way ANOVA for continuous variables and chi-squared for categorical variables

### Supplementary Table 2 - list of questionnaire categories

| **Questionnaire category** | **Number of questions** |
| --- | --- |
| Personal history | 181 |
| Family history | 86 |
| Sleep | 71 |
| Nutrition | 35 |
| Stree | 34 |
| Pollution, environment | 21 |
| Personal details | 18 |
| Skin health | 17 |
| Mood | 15 |
| Sexual health | 14 |
| Sport habits | 12 |
| Hair & nails health | 9 |

### Supplementary Table 3 - list of health measurements, with corresponding code in SNOMED/LOINC systems, if available

| **Name** | **LOINC** | **SNOMED** |
| --- | --- | --- |
| Antioxidant power.measured [Molar/volume] (Serum) |  |  |
| Neutrophils [#/volume] (Blood) by Automated count | 751-8 |  |
| Leukocytes [Presence] (Urine) by Test strip | 20408-1 |  |
| Cortisol --8AM specimen/Cortisol -- 8PM specimen [Molar Ratio] (Saliva) |  |  |
| Folate [Mass/volume] (Red Blood Cells) | 2283-0 |  |
| Prostate specific Ag [Mass/volume] (Serum) | 2857-1 |  |
| Pecan or Hickory Nut IgG Ab [Mass/volume] (Serum) | 63108-5 |  |
| Retinol [Mass/volume] (Serum) | 2923-1 |  |
| Android fat mass/Body tissue mass [Mass Ratio] |  |  |
| Omega 3 Index (Serum) | 88998-0 |  |
| Salmon IgG Ab [Mass/volume] (Serum) | 60400-9 |  |
| Epstein Barr virus nuclear IgG Ab [Units/volume] (Serum) | 31374-2 |  |
| Human immunodeficiency viruses (HIV) 1+2 [Presence] (Serum) | 7918-6 |  |
| Adaptation resource |  |  |
| C reactive protein [Mass/volume] (Serum) | 1988-5 |  |
| Testosterone [Mass/volume] (Serum) | 2986-8 |  |
| Alanine aminotransferase [Enzymatic activity/volume] (Serum) | 1742-6 |  |
| Cortisol [Moles/volume] (Saliva) --8 PM specimen | 58677-6 |  |
| Sex hormone binding globulin [Moles/volume] (Serum) | 13967-5 |  |
| Vanillylmandelate [Mass/volume] (24 hour Urine) | 26706-2 |  |
| Albumin [Mass/volume] (Serum) by Bromocresol purple (BCP) dye binding method | 61152-5 |  |
| Methyl acetate delta as measured in exhaled gas minus calibrated in fresh air |  |  |
| Banana IgG Ab [Mass/volume] (Serum) | 60352-2 |  |
| Dihomo-gamma-linoleneate/Fatty acids.total [Mass Ratio] (Serum) |  |  |
| Folate [Mass/volume] (Serum) | 2284-8 |  |
| Arachidonate/Eicosapentaenoate [Mass Ratio] (Serum) |  |  |
| Gamma linolenate/Fatty acids.total [Mass Ratio] (Serum) |  |  |
| pH (Urine) by Test strip | 5803-2 |  |
| Soluble urokinase plasminogen activator receptor (suPAR) [Mass/volume] (Serum) | 101170-9 |  |
| Forced vital capacity (FVC) [Z-score] |  |  |
| Whole Egg IgG Ab [Mass/volume] (Serum) | 45201-1 |  |
| Transferrrin/Iron saturation [Mass Ratio] (Serum) |  |  |
| 3-Methoxy-4-Hydroxyphenylglycol/Creatinine [Mass Ratio] (Urine) |  |  |
| Hydrogen delta as measured in exhaled gas minus calibrated in fresh air |  |  |
| 3,4-Dihydroxyphenylacetate [Mass/volume] (Urine) |  |  |
| Coenzyme Q10 [Mass/volume] (Serum) | 27923-2 |  |
| Hearing right ear - 4000 Hz | 89023-6 |  |
| Selenium [Mass/volume] (Serum) | 5724-0 |  |
| Eicosapentaenoate/Fatty acids.total [Mass Ratio] (Serum) |  |  |
| Average grip strength (left hand) | 83173-5 |  |
| Serotonin [Mass/volume] (24 hour Urine) | 56978-0 |  |
| Oleate/Fatty acids.total [Mass Ratio] (Serum) |  |  |
| Insulin sensitivity index (Serum) by calculation | 47214-2 |  |
| Forced expiratory volume in 1 second (FEV1) / Forced vital capacity (FVC) [Volume Ratio] [Z-score] |  |  |
| Cholesterol.total/Cholesterol in HDL [Molar Ratio] (Serum) | 32309-7 |  |
| Garlic IgG Ab [Mass/volume] (Serum) | 60374-6 |  |
| Hearing left ear - 1000 Hz | 89016-0 |  |
| Cow whey IgG Ab [Mass/volume] (Serum) | 59058-8 |  |
| Homeostasis model assessment (Serum) | 47214-2 |  |
| Cortisol [Moles/volume] (Saliva) --8 AM specimen | 58675-0 |  |
| Aspartate aminotransferase [Enzymatic activity/volume] (Serum) | 1920-8 |  |
| Hepatitis B virus surface Ag [Units/volume] in Serum | 58452-4 |  |
| Homovanillate/5-Hydroxyindoleacetate [Mass Ratio] (Urine) |  |  |
| Chicken meat IgG Ab [Mass/volume] (Serum) | 99366-7 |  |
| Baevsky stress index |  |  |
| Beta globulin [Mass/volume] (Serum) by Electrophoresis | 2871-2 |  |
| Nitric oxide measured in fresh air for calibration |  |  |
| Alpha tocopherol [Mass/volume] (Serum) | 1823-4 |  |
| Vanillylmandelate/Creatinine [Mass Ratio] (Urine) | 3124-5 |  |
| Hepatitis C virus Ab [Units/volume] in Serum | 22327-1 |  |
| Maximum grip strength (right hand) |  |  |
| Glutathione peroxidase [Enzymatic activity/volume] (Blood) | 45328-2 |  |
| Eosinophils [#/volume] (Blood) by Automated count | 711-2 |  |
| Bilirubin.conjugated [Mass/volume] (Serum) | 15152-2 |  |
| Sample hemolyzed [Presence] (Serum) | 20393-5 |  |
| Follitropin [Units/volume] (Serum) | 15067-2 |  |
| DOPamine/Creatinine [Mass Ratio] (Urine) | 13733-1 |  |
| Omega 3/Fatty acids.total [Mass Ratio] (Serum) |  |  |
| Hearing left ear - 500 Hz | 89024-4 |  |
| Glutathione peroxidase [Enzymatic activity/mass] (Red Blood Cells) | 45378-7 |  |
| Gamma glutamyl transferase [Enzymatic activity/volume] (Serum) | 2324-2 |  |
| Bone density [Z-score] |  |  |
| Cobalamin (Vitamin B12) [Mass/volume] (Serum) | 2132-9 |  |
| Vagal Index |  |  |
| Monocytes [#/volume] (Blood) by Automated count | 742-7 |  |
| Homovanillate/Creatinine [Mass Ratio] (Urine) | 11146-8 |  |
| Cholesterol non HDL [Mass/volume] (Serum) | 43396-1 |  |
| CLA3 C18:2(9c,12t)/Fatty acids.total [Mass Ratio] (Serum) |  |  |
| Hearing right ear - 500 Hz | 89025-1 |  |
| Volume of 24 hour Urine | 3167-4 |  |
| Triiodothyronine (T3) [Mass/volume] (Urine) |  |  |
| 3-Methoxy-4-Hydroxyphenylglycol [Mass/volume] (24 hour Urine) | 21043-5 |  |
| Glucose [Presence] (Urine) by Test strip | 25428-4 |  |
| Resting heart rate | 40443-4 | 444981005 |
| Body lean mass | 91557-9 |  |
| Superoxide dismutase [Enzymatic activity/volume] (Blood) | 45333-2 |  |
| Herpes simplex virus 1+2 IgG Ab [Units/volume] in Serum by Immunoassay | 27948-9 |  |
| Elaidic/Fatty acids.total [Mass Ratio] (Serum) |  |  |
| Body fat mass/Body lean mass [Mass Ratio] [Z-score] |  |  |
| Copper/Zinc [Mass Ratio] (Serum) | 48767-8 |  |
| CLA2 C18:2(10t,12c)/Fatty acids.total [Mass Ratio] (Serum) |  |  |
| Cholesterol in HDL [Mass/volume] (Serum) | 2085-9 |  |
| Bone density [T-score] |  |  |
| Protein [Mass/volume] (Serum) | 2885-2 |  |
| Iodine [Mass/time] (24 hour Urine) | 2492-7 |  |
| Triglyceride [Mass/volume] (Serum) | 2571-8 |  |
| Zinc [Mass/volume] (Serum) | 5763-8 |  |
| Hematocrit [Volume Fraction] (Blood) by Automated count | 4544-3 |  |
| SpO2 | 59408-5 | 431314004 |
| Thyroxine (T4) free [Moles/volume] (Serum) | 14920-3 |  |
| Dehydroepiandrosterone (DHEA) [Moles/volume] (Saliva) --8PM specimen | 74361-7 |  |
| Superoxide dismutase [Enzymatic activity/mass] (Red Blood Cells) | 56479-9 |  |
| Hearing left ear - 2000 Hz | 89018-6 |  |
| Sample icteric [Presence] (Serum) | 20392-7 |  |
| Cholesterol in LDL [Mass/volume] (Serum) by Calculation | 13457-7 |  |
| Trans-palmitoleate/Fatty acids.total [Mass Ratio] (Serum) |  |  |
| Insulin [Moles/volume] (Serum) | 14796-7 |  |
| Prostate Specific Ag Free [Mass/volume] (Serum) | 10886-0 |  |
| Cholesterol [Mass/volume] (Serum) | 2093-3 |  |
| Potassium [Moles/volume] (Serum) | 2823-3 |  |
| Forced expiratory volume in 1 second (FEV1) pre-bronchodilator | 20157-4 |  |
| Stearic acid/Fatty acids.total [Mass Ratio] (Serum) |  |  |
| Serotonin/Creatinine [Mass Ratio] (Urine) |  |  |
| Linoleate/Dihomo-gamma-linoleneate [Mass Ratio] (Serum) |  |  |
| Alpha tocopherol/Cholesterol [Mass Ratio] (Serum) |  |  |
| Potato IgG Ab [Mass/volume] (Serum) | 35544-6 |  |
| Alpha 1 globulin [Mass/volume] (Serum) by Electrophoresis | 2865-4 |  |
| Sample lipemic [Presence] (Serum) | 20394-3 |  |
| Casein IgG Ab [Mass/volume] (Serum) | 53829-8 |  |
| Mercury [Mass/volume] (Serum) | 5687-9 |  |
| Average grip strength (hands asymmetry) |  |  |
| Glucose [Mass/volume] (Serum) --pre-meal | 53049-3 |  |
| Prostate Specific Ag Free/Prostate specific Ag.total [Mass Ratio] (Serum) | 12841-3 |  |
| Hyaluronate [Mass/volume] (Serum) | 12736-5 |  |
| Alpha-linolenate/Fatty acids.total [Mass Ratio] (Serum) |  |  |
| Lactalbumin alpha IgG Ab [Mass/volume] (Serum) | 63418-8 |  |
| Linoleate/Fatty acids.total [Mass Ratio] (Serum) |  |  |
| Heart rate variability (HRV) Index |  |  |
| Chloride [Moles/volume] (Serum) | 2075-0 |  |
| Platelets [#/volume] (Blood) by Automated count | 777-3 |  |
| Palmitoleate/Fatty acids.total [Mass Ratio] (Serum) |  |  |
| Testosterone.free+weakly bound/Testosterone.total [Mass Ratio] (Serum) | 6891-6 |  |
| Cocoa IgG Ab [Mass/volume] (Serum) | 99367-5 |  |
| Gamma globulin/Protein.total [Mass Ratio] (Serum) by Electrophoresis | 13983-2 |  |
| LDL.oxidized Ab [Units/volume] (Serum) | 48143-2 |  |
| Alpha 2 globulin [Mass/volume] (Serum) by Electrophoresis | 2868-8 |  |
| Pregnenolone [Mass/volume] (Serum) | 2837-3 |  |
| Hepatitis B virus surface Ab [Units/volume] in Serum | 16935-9 |  |
| Iodine [Mass/volume] (24 hour Urine) | 26842-5 |  |
| Arsenic [Mass/volume] (Serum) | 5585-5 |  |
| Android fat mass/Gynoid fat mass [Mass Ratio] |  |  |
| Transferrin [Mass/volume] (Serum) | 3034-6 |  |
| Bilirubin.total [Mass/volume] (Serum) | 1975-2 |  |
| Wheat IgG Ab [Mass/volume] (Serum) | 35537-0 |  |
| Gynoid fat mass/Body tissue mass [Mass Ratio] |  |  |
| Myristate/Fatty acids.total [Mass Ratio] (Serum) |  |  |
| Sodium [Moles/volume] (Serum) | 2951-2 |  |
| Homocysteine [Moles/volume] (Serum) | 13965-9 |  |
| Egg white IgG Ab [Mass/volume] (Serum) | 35535-4 |  |
| Glomerular filtration rate/1.73 sq M.predicted [Volume Rate/Area] (Serum) by Creatinine-based formula (CKD-EPI) | 62238-1 |  |
| 5-Hydroxyindoleacetate/Creatinine [Mass Ratio] (Urine) | 11145-0 |  |
| Lymphocytes [#/volume] (Blood) by Automated count | 731-0 |  |
| Average grip strength (right hand) |  |  |
| Leukocytes [#/volume] (Blood) by Automated count | 6690-2 |  |
| Eosinophils/100 leukocytes (Blood) by Automated count | 713-8 |  |
| Forced expiratory volume in 1 second (FEV1) / Forced vital capacity (FVC) [Volume Ratio] pre-bronchodilator | 19926-5 |  |
| Testosterone Free [Mass/volume] (Serum) | 2991-8 |  |
| Apolipoprotein A-I/Apolipoprotein B [Mass Ratio] (Serum) | 13462-7 |  |
| Prolactin [Mass/volume] (Serum) | 2842-3 |  |
| Lipoprotein a [Mass/volume] (Serum) | 10835-7 |  |
| EPINEPHrine [Mass/volume] (24 hour Urine) | 32015-0 |  |
| Neutrophils/100 leukocytes (Blood) by Automated count | 770-8 |  |
| Hydrogen sulfide delta as measured in exhaled gas minus calibrated in fresh air |  |  |
| Eicosatrienoic acid/Fatty acids.total [Mass Ratio] (Serum) |  |  |
| Platelet mean volume [Entitic volume] (Blood) | 28542-9 |  |
| Biological age |  |  |
| Hearing right ear - 1000 Hz | 89017-8 |  |
| Antioxidant power.total [Molar/volume] (Serum) | 55219-0 |  |
| Soybean IgG Ab [Mass/volume] (Serum) | 35546-1 |  |
| MCH [Entitic mass] (Blood) by Automated count | 785-6 |  |
| Glucose mean value [Mass/volume] (Blood) estimated from Glycated hemoglobin | 27353-2 |  |
| Free/total testosterone ratio (Serum) | 15432-8 |  |
| Hydrogen in fresh air for calibration |  |  |
| Automonic balance |  |  |
| Triiodothyronine (T3) Free [Moles/volume] (Serum) | 14928-6 |  |
| Basophils [#/volume] (Blood) by Automated count | 704-7 |  |
| Gamma globulin [Mass/volume] (Serum) by Electrophoresis | 2874-6 |  |
| Erythrocytes [#/volume] (Blood) by Automated count | 789-8 |  |
| EPINEPHrine/Creatinine [Mass Ratio] (24 hour Urine) | 43248-4 |  |
| Testosterone.bound/Testosterone.total (Serum) |  |  |
| Alkaline phosphatase [Enzymatic activity/volume] (Serum) | 6768-6 |  |
| HPV (16, 18, 31, 33, 35, 39, 45, 51, 52, 56, 58, 59, 66 & 68) DNA [Presence] by NAA with Probe detection (Vaginal specimen) by Swab | 77378-8 |  |
| Nickel [Mass/volume] (Serum) | 5702-6 |  |
| 3,4-Dihydroxyphenylacetate/Creatinine [Mass Ratio] (Urine) |  |  |
| Cod IgG Ab [Mass/volume] (Serum) |  |  |
| Docosahexaenoate/Fatty acids.total [Mass Ratio] (Serum) |  |  |
| Arachidonate/Fatty acids.total [Mass Ratio] (Serum) |  |  |
| Monocytes/100 leukocytes (Blood) by Automated count | 5905-5 |  |
| Methyl acetate in fresh air for calibration |  |  |
| Goat milk IgG Ab [Mass/volume] (Serum) | 63133-3 |  |
| Copper [Mass/volume] (Serum) | 5631-7 |  |
| Body tissue mass |  |  |
| Hydrogen measured in exhaled gas |  |  |
| Thyrotropin [Units/volume] (Serum) | 3016-3 |  |
| Trans-vaccenate/Fatty acids.total [Mass Ratio] (Serum) |  |  |
| Insulin-like growth factor-I [Mass/volume] (Serum) | 2484-4 |  |
| Hearing left ear - 4000 Hz | 89022-8 |  |
| Onion IgG Ab [Mass/volume] (Serum) | 35541-2 |  |
| Iron saturation [Mass Fraction] (Serum) | 2502-3 |  |
| Cortisol [Moles/volume] (Saliva) --8 AM + 30' specimen |  |  |
| Hemoglobin A1c/Hemoglobin.total (Blood) | 4548-4 |  |
| Manganese [Mass/volume] (Serum) | 5683-8 |  |
| Adaptation index |  |  |
| MCV [Entitic volume] (Blood) by Automated count | 787-2 |  |
| Maximum grip strength (hands asymmetry) |  |  |
| Antioxidant power.calculated [Molar/volume] (Serum) |  |  |
| Basophils/100 leukocytes (Blood) by Automated count | 706-2 |  |
| Forced vital capacity (FVC) pre-bronchodilator | 19877-0 |  |
| Cystatin C [Mass/volume] (Serum) | 33863-2 |  |
| Fluorescence of Advanced Glycation Endproducts (AGEs) |  |  |
| Dehydroepiandrosterone sulfate (DHEA-S) [Mass/volume] (Serum) | 2191-5 |  |
| Hearing right ear - 2000 Hz | 89019-4 |  |
| Iron binding capacity [Mass/volume] (Serum) | 2500-7 |  |
| 8-Hydroxydeoxyguanosine [Mass/volume] (Serum) |  |  |
| DOPamine [Mass/volume] (Urine) | 2217-8 |  |
| MCHC [Mass/volume] (Blood) by Automated count | 786-4 |  |
| High-sensitivity C-reactive protein (hs-CRP) [Mass/volume] (Serum) | 71426-1 |  |
| Bilirubin.indirect [Mass/volume] (Serum) | 1971-1 |  |
| Creatinine [Mass/volume] (Urine) | 2161-8 |  |
| Hemoglobin [Mass/volume] (Blood) | 718-7 |  |
| Hydrogen sulfide measured in exhaled gas |  |  |
| Norepinephrine/Creatinine [Mass Ratio] (24 hour Urine) | 44341-6 |  |
| Omega 6/Omega 3[Mass Ratio] (Blood) | 90910-1 |  |
| Body mass |  |  |
| Adaptation strain |  |  |
| Hepatitis B virus core Ab [Presence] in Serum | 16933-4 |  |
| Apolipoprotein B [Mass/volume] (Serum) | 1884-6 |  |
| Shrimp IgG Ab [Mass/volume] (Serum) | 60403-3 |  |
| Ferritin [Mass/volume] (Serum) | 2276-4 |  |
| Omega 6/Fatty acids.total [Mass Ratio] (Serum) |  |  |
| Testosterone.free+weakly bound [Moles/volume] (Serum) | 41018-3 |  |
| Heart rate variability (HRV) pNN50 |  |  |
| Homovanillate [Mass/volume] (24 hour Urine) | 53595-5 |  |
| Corn IgG Ab [Mass/volume] (Serum) | 99467-3 |  |
| Maximum grip strength (left hand) |  |  |
| Tomato IgG Ab [Mass/volume] (Serum) | 35547-9 |  |
| Cortisol/DHEA [Molar Ratio] (Saliva) |  |  |
| Pork IgG Ab [Mass/volume] (Serum) | 35543-8 |  |
| Protein [Presence] (Urine) by Test strip | 20454-5 |  |
| Methyl acetate measured in exhaled gas |  |  |
| Nitric oxide measured in exhaled gas | 74369-0 |  |
| Urate [Mass/volume] (Serum) | 3084-1 |  |
| Iron [Mass/volume] (Serum) | 2498-4 |  |
| Vaccenate/Fatty acids.total [Mass Ratio] (Serum) |  |  |
| Aging rate |  |  |
| Lymphocytes/100 leukocytes (Blood) by Automated count | 736-9 |  |
| Nitric oxide delta as measured in exhaled gas minus calibrated in fresh air |  |  |
| Forced expiratory volume in 1 second (FEV1) [Z-score] |  |  |
| Hydrogen sulfide in fresh air for calibration |  |  |
| Gluten IgG Ab [Mass/volume] (Serum) | 63091-3 |  |
| Hemoglobin A1c/Hemoglobin.total (Blood) by IFCC protocol | 59261-8 |  |
| Palmitate/Fatty acids.total [Mass Ratio] (Serum) |  |  |
| Apolipoprotein A-I [Mass/volume] (Serum) | 1869-7 |  |
| Brewer's yeast IgG Ab [Mass/volume] (Serum) | 60413-2 |  |
| Creatinine [Mass/volume] (Serum) | 2160-0 |  |
| Blood [Presence] in Urine by Test strip |  |  |
| Triiodothyronine (T3) [Moles/time] (24 hour Urine) | 97165-5 |  |
| Beta lactoglobulin IgG Ab [Mass/volume] (Serum) | 63090-5 |  |
| Visceral adipose tissue (VAT) mass |  |  |
| 8-Hydroxydeoxyguanosine/Creatinine [Mass Ratio] (Serum) |  |  |
| 5-Hydroxyindoleacetate [Mass/volume] (24 hour Urine) | 31203-3 |  |
| Cytomegalovirus IgG Ab [Units/volume] (Serum) | 7852-7 |  |
| Chlamydia trachomatis IgG Ab [Units/volume] (Serum) by Immunoassay | 26715-3 |  |
| 25-hydroxyvitamin D3+D2 [Mass/volume] (Serum) | 62292-8 |  |
| Cortisol Awakening Response (Saliva) |  |  |
| Tuna IgG Ab [Mass/volume] (Serum) | 60410-8 |  |
| Rumenate/Fatty acids.total [Mass Ratio] (Serum) |  |  |
| Body fat mass |  |  |
| Norepinephrine [Mass/volume] (24 hour Urine) | 27221-1 |  |
| Epstein Barr virus capsid IgG Ab [Units/volume] (Serum) | 7885-7 |  |
| Body temperature | 8310-5 | 386725007 |
| Neck circumference | 56074-8 | 420236003 |
| Waist circumference | 56115-9 | 1162535003 |
| Heart rate while sitting | 69000-8 | 364075005 |
| Systolic blood pressure while sitting | 8459-0 | 407554009 |
| Weight | 29463-7 | 27113001 |
| Height | 8302-2 | 248333004 |
| Diastolic blood pressure while sitting | 8453-3 | 407555005 |

### Supplementary Table 4 - The STANDING Together consensus recommendations for the documentation of Health Datasets

| **Recommendation** | **Detail** |
| --- | --- |
| 1.1a: dataset summary | The cohort was designed to provide a comprehensive and longitudinal understanding of health determinants, allowing for potential long-term follow-up. It was established to comprehensively diagnose diseases, assess early risk markers, and prevent future disease occurrences, in a health center focused on personalized prevention. Participants included in the cohort were adults aged 18 years or older, enrolled in the Zoī program either as paying customers or as beneficiaries of a company-sponsored program. |
| 1.1b: dataset identity and access | Access to the Zoī cohort is possible for research purposes, and can only be provided at no cost to investigators, ensuring that financial barriers do not hinder scientific progress.  Hence, collaboration with Zoī is guided by strict adherence to legal and ethical standards, prioritizing data confidentiality and security. Proposals for research projects involving the Zoī cohort are to be reviewed by the Zoī Scientific and Medical Committee that evaluates missions based on scientific merit, and an independent Ethical Committee for ethical considerations. |
| 1.1c: reasons behind dataset creation and its purpose(s) | The dataset collected through this cohort enables the investigation of the complex relationships between biological, behavioral, and environmental factors and their influence on health outcomes. Additionally, by analyzing adherence to personalized recommendations, this cohort offers a unique opportunity to study the effectiveness of preventive strategies over time. Moreover, the cohort offers valuable opportunities for machine learning applications in the health domain. |
| 1.1d: data origin | Participants included in the cohort were adults aged 18 years or older, enrolled in the Zoī program either as paying customers or as beneficiaries of a company-sponsored program. All participants were informed generally and individually prior to inclusion, with the possibility to object, in accordance with national regulations. |
| 1.1e: data sampling and aggregation from multiple sources | The dataset isn’t aggregated from multiple sources, as it is based only from the Zoī cohort. Dataset documentation and standardisation uses both the LOINC and SNOMED CT collections. |
| 1.1f: data shifts over time | The population included in the Zoī cohort isn’t expected to change in the near future. Medical practice and devices used are documented in detail, and any changes will be apparent.  Additional clinical prediction models may be added to the protocol, but they may be retrospectively estimated on the whole cohort. |
| 1.2a: composition of groups within dataset | Participants included in the cohort were enrolled in the Zoī program either as paying customers or as beneficiaries of a company-sponsored program. Those two groups may differ in preventive health engagement. |
| 1.2b: recording of individuals' attributes | All individuals’ attributes are either from self-reported questionnaires, or from the subsequent measures during the check-up: clinical examinations, medical devices (electrocardiogram, spirometry, blood pressure), laboratory tests (urine, blood, saliva), and medical imaging. Validated clinical prediction models were retrospectively applied to the cohort data, focusing on disease prevention.  Most diagnoses were automatically calculated from clinical and biological measurements using internationally recognized criteria and validated by medical doctors. More complex diagnoses were generated by MDs and extracted from medical records. In cases of missing diagnostic reports, retrospective assessments were conducted by trained clinicians.  Attributes missing or recorded as “prefer not to say” are described as such (Table 1) and not imputed. |
| 1.2c: groups at risk of disparate health outcomes | Participants included in the cohort were enrolled in the Zoī program either as paying customers or as beneficiaries of a company-sponsored program. Those two groups may differ in preventive health engagement, and thus in health outcomes. |
| 1.3a: limitations of the dataset | Participants in this cohort are predominantly highly educated; they exhibit a lower prevalence of smoking, obesity, and better self-rated health compared to the general population. While it limits the generalizability of findings to the broader population, it provides a focused opportunity to study health dynamics in a group that is assumed to be well-informed and proactive about health. |
| 1.3b: modifications made to the data | None of the data is synthetic nor imputed.  Validated clinical prediction models were retrospectively applied to the cohort data. |
| 1.3c: missing data | Missing data may occur in the future if the check-up protocol is modified, from added or deleted clinical examinations, medical devices, laboratory tests or medical imaging. This will lead to systematic differences between older and newer participants' data. Currently, attributes missing or recorded as “prefer not to say” are described as such (Table 1) and not imputed. |
| 1.3d: known or potential bias caused or exacerbated by data acquisition and processing | Standardized protocols were implemented across all clinical assessments to ensure consistency and reliability. Measurements are performed using the same equipment, with the same team of HCPs, under controlled environmental conditions, including lighting, sound, temperature and humidity of each room. Data collection was almost entirely automated, minimizing the risk of entry errors and missing values. As a result, no known bias may be caused or exacerbated by data acquisition or processing. |
| 1.3e: known or potential exclusion introduced by data collection | All participants had the opportunity to object to inclusion, in accordance with national regulations. Participants who objected were excluded and the data excluded, removing any possibility of attempting to mitigate this bias. |
| 1.3f: known or potential bias in assigned or derived labels | Most diagnoses were automatically calculated from clinical and biological measurements using internationally recognized criteria and validated by medical doctors. As a consequence, the set of labels (diagnoses) to include is fixed and predetermined, focused on simple diagnoses.  However, to mitigate this bias, more complex diagnoses were generated by MDs at check-up time and extracted from medical records. In cases of missing diagnostic reports, retrospective assessments were conducted by trained clinicians. |
| 1.4a: ethics and governance | Institutional review board approval was obtained for this study. All participants were informed generally and individually prior to inclusion, with the possibility to object, in accordance with national regulations.  No individual-identifiable data or images are included in this manuscript. The study involved only retrospective, pseudonymized data.  For free-text data, pseudonymization is performed using a machine learning approach based on a standard token classification model for general-purpose named entity recognition (NER), following a framework published elsewhere (Tannier & al, Methods Inf Med. 2024) |
| 1.4b: patient and public participation | The protocol is reviewed by the Zoī Scientific and Medical Committee, as described in <https://www.zoi.com/en/medical-committee>.  No patient nor public participation groups were involved. |
| 1.4c: bias and impact assessments | No formal assessment of bias or societal impact has been previously conducted. |

### Supplementary Fig.1 - Distribution of some specific health scores calculated at check-up time: (A) SCORE2 for cardiovascular event risk,(B) aMAP for liver cancer risk, (C) LLPv3 for lung cancer risk.

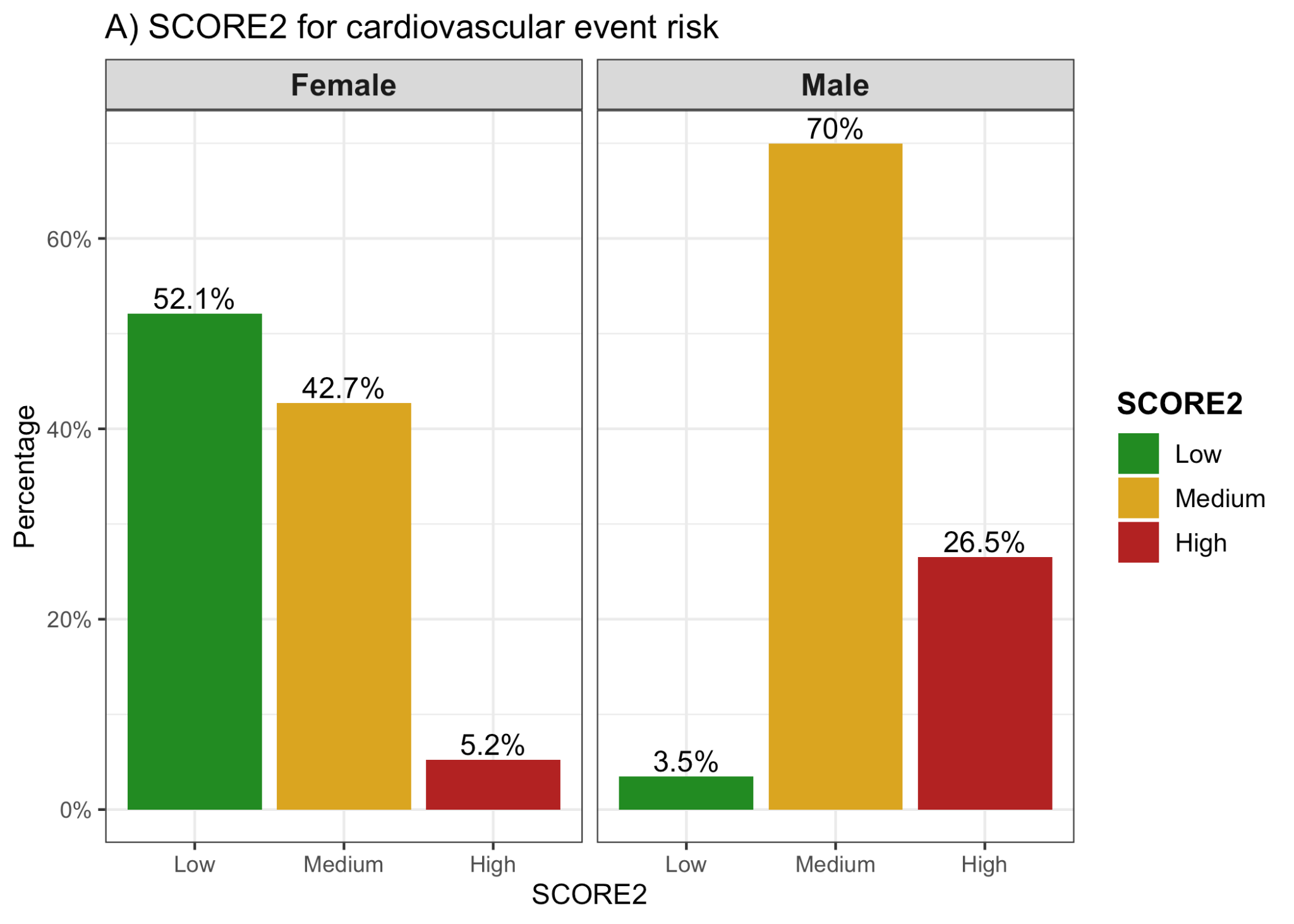

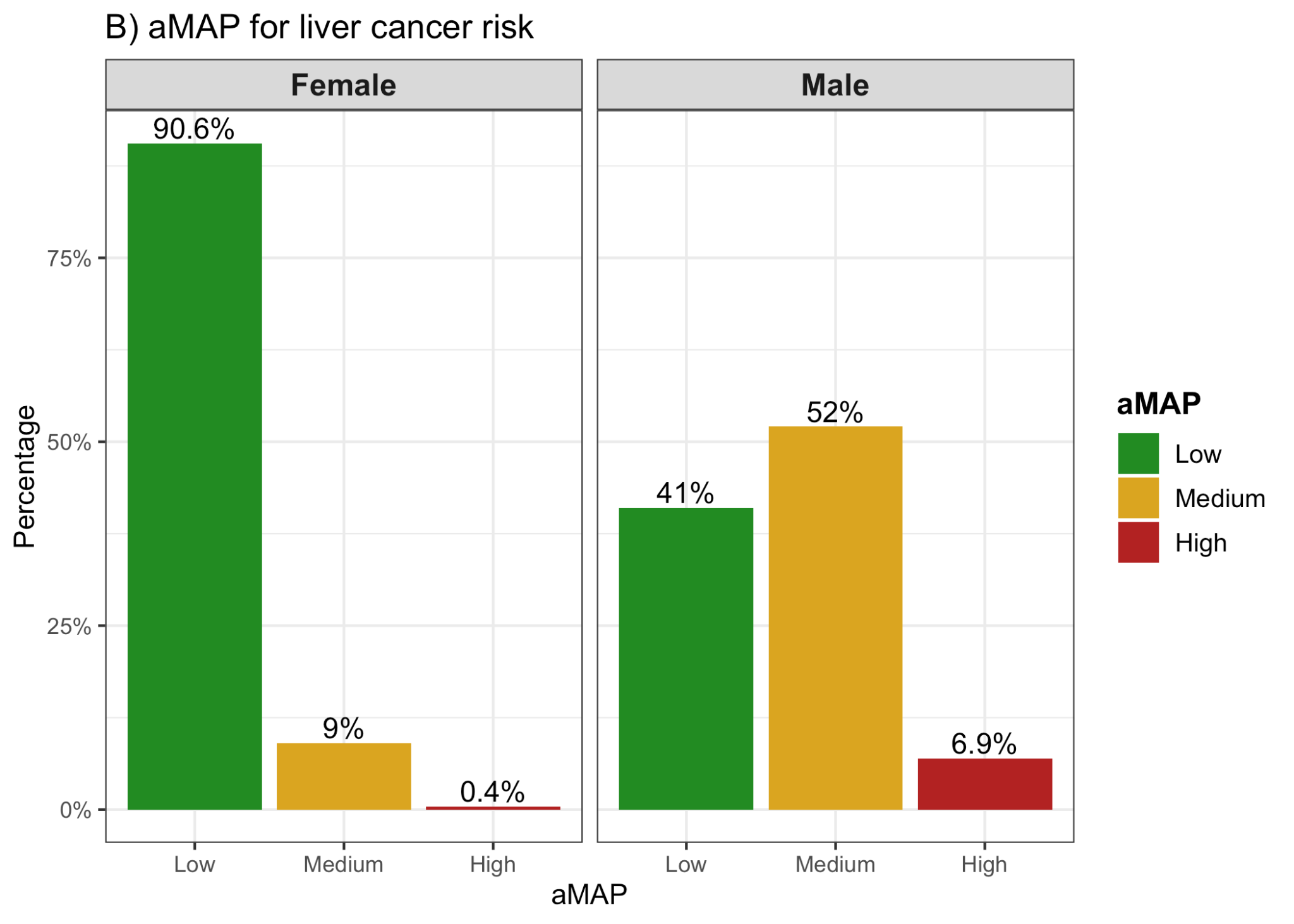

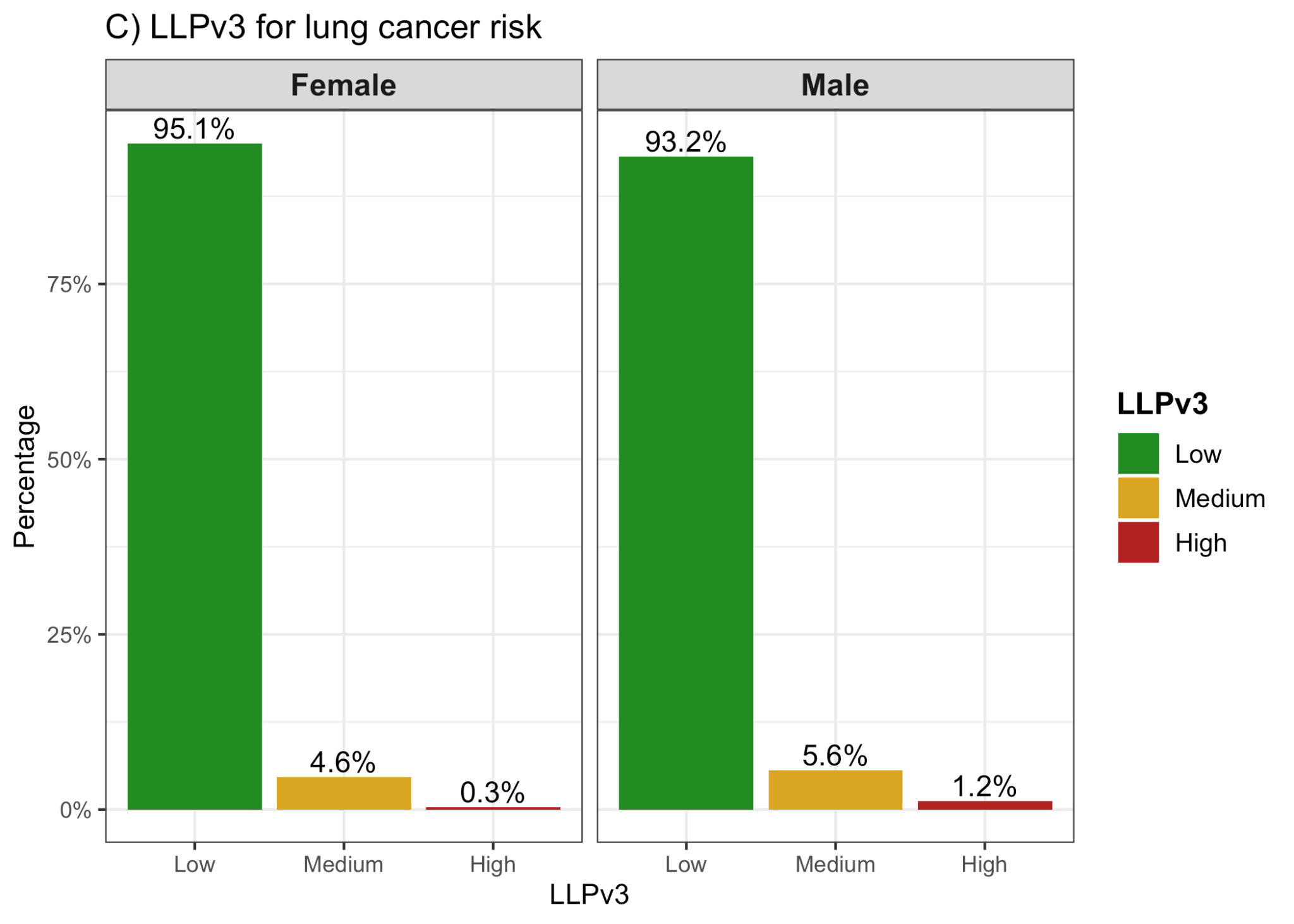

### Supplementary Fig.2 - Example of DEXA report

**Please reach out to the author if you'd like a copy.**

### Supplementary Fig.3 - Example of spirometry report

**Please reach out to the author if you'd like a copy.**

### Supplementary Fig.4 - Example of grip strength report

**Please reach out to the author if you'd like a copy.**

### Supplementary Fig.5 - Example of OCT image

**Please reach out to the author if you'd like a copy.**
